# A foundational neuronal protein network model unifying multimodal genetic, transcriptional, and proteomic perturbations in schizophrenia

**DOI:** 10.1101/2025.05.02.25326757

**Authors:** Greta Pintacuda, Yu-Han H. Hsu, Petra Páleníková, Ugne Dubonyte, Nadine Fornelos, Miao Chen, Daya Mena, Julia C. Biagini, Travis Botts, Makayla Martorana, Danzel Rebelo, Joshua K.T. Ching, Ethan Crouse, Hilena Gebre, Xian Adiconis, Nathan Haywood, Sean Simmons, Michel Weïwer, Derek Hawes, Olli Pietilainen, Thomas Werge, Ka Wan Li, August B. Smit, Agnete Kirkeby, Joshua Z. Levin, Ralda Nehme, Kasper Lage

## Abstract

Schizophrenia (SCZ) is a complex psychiatric disorder with a diverse genetic landscape, involving common regulatory variants, rare protein-coding mutations, structural genomic rearrangements, and transcriptional dysregulation. A critical challenge in developing rationally designed therapeutics is understanding how these various factors converge to disrupt cellular networks in the human brain, ultimately contributing to SCZ. Towards this aim, we generated multimodal data, including SCZ-specific protein-protein interactions in stem-cell-derived neuronal models and adult postmortem cortex, integrated with genetic and transcriptomic datasets from individuals with psychiatric disorders. We identified three distinct neuron-specific SCZ protein networks, or modules, significantly enriched for genetic and transcriptional perturbations associated with SCZ. The relevance of these modules was validated through whole-cell proteomics in patient-derived neurons, revealing their disruption in 22q11.2 deletion carriers diagnosed with SCZ. We demonstrated their therapeutic potential by showing that these modules are targets of GSK3 inhibition using phosphoproteomics. Our findings present a foundational model that integrates genetic, transcriptional, and proteomic perturbations in SCZ. This model provides a cohesive framework for understanding how polygenic and multimodal perturbations affect neuronal pathways in the human brain, as well as a data-driven pathway resource for identifying potential drug targets to reverse disruptions observed in these neuronal networks.

## Introduction

Schizophrenia (SCZ) is a complex psychiatric disorder with a significant global prevalence and a highly polygenic genetic architecture [1], shaped by common risk alleles [2], rare protein-coding mutations [3, 4], and structural genomic rearrangements such as copy number variants (CNV) [5] affecting hundreds of genes [6]. Additionally, transcriptional differences at the single-cell level have been observed in postmortem brains of individuals with SCZ compared to controls, regardless of the genetic background [7]. These studies both highlight the genetic complexity of the disorder and point to excitatory and inhibitory neurons in the prefrontal cortex as two major cell types that not only concentrate polygenic risk, but also exhibit significant transcriptional changes linked to SCZ. This suggests that a foundational framework for understanding how genetic, transcriptional, and protein-level perturbations converge in molecular pathways and networks in these cell types will be critical to understanding and reversing SCZ pathology in the human brain.

Despite these discoveries, how cell-type-specific effects driven by the hundreds of SCZ-associated variants align with transcriptional changes, and how perturbations at the genomic, transcriptomic, and proteomic levels converge on the mechanistic pathways that drive SCZ remain poorly understood. This gap hinders the development of therapeutic strategies, with most efforts still focused on symptom management and the acute treatment of psychotic episodes [8, 9]. A major challenge for the coming decade will be to integrate vast multimodal datasets in psychiatry—such as those from the PsychENCODE Consortium [10] and the Human BRAIN Initiative [11]—into a cohesive mechanistic model of disease. Such a model of SCZ would aim to explain how genetic and transcriptional perturbations of hundreds of genes assemble into cell-type-specific pathways that contribute to disease, while also providing a validation strategy in patient-derived neurons to support causality. This foundational model could not only bridge a gap to advance SCZ research, but also provide a framework for integrating biological insights with rational drug discovery efforts. Indeed, this framework could extend to other neurodevelopmental and neuropsychiatric disorders with similar genetic complexity to SCZ, highlighting its broader applicability.

One promising strategy to address this challenge is to leverage stem cell technologies to model brain cell types most relevant to SCZ, enabling the mapping of cell-type-specific pathways and the direct testing of mechanistic hypotheses in comparable neuronal cellular models derived from individuals with SCZ. Although these cellular models have well-recognized limitations, such as incomplete maturation [12] and limited capacity to replicate cell-to-cell interactions within complex tissues [13], they are increasingly valuable for multimodal functional analysis. This includes analyzing the biology of non-SCZ linked neurons as a background and using this background to compare to perturbations observed in cells derived from individuals with highly penetrant SCZ protein-coding mutations [14] or CNVs [5]. These models also provide a robust platform for evaluating cellular responses to drug treatments [15]. If SCZ-relevant pathways identified in cell models are both shown to reflect the biology and biochemistry of complex adult human brain tissue and are validated in neurons derived from individuals with SCZ, they could yield critical insights into SCZ pathophysiology and inform the development of targeted therapeutics.

Previous work by our group and others have demonstrated that combining stem cell modeling with interaction proteomics is a powerful approach for identifying convergent mechanisms in neurodevelopmental, neuropsychiatric, and other complex disorders [16–19]. Here, we further evolve this strategy to identify neuron-specific protein pathways linked to SCZ at the genetic, transcriptional, and proteomic levels. We leveraged the non-stoichiometric nature of interactions mapped by protein-protein interaction (PPI) networks—including second- and third-degree connections as well as dynamic interactions—to construct a comprehensive network of protein pathway relationships anchored on 13 proteins with rare, penetrant variants strongly associated with SCZ risk. These pathway relationships were identified through immunoprecipitation-mass spectrometry (IP-MS) in both stem cell-derived neuronal models and postmortem adult cortical tissue. Following rigorous quality control and data harmonization, we confirmed that the PPI data encompassed true biological pathways despite the *in vitro* nature of the cell models and technical variability inherent to IP-MS. Moreover, many pathway relationships observed in the neuronal models were also found in the postmortem cortex, despite differences in cellular complexity and developmental age. These findings validate that our approach is effective for mapping bona fide protein pathways in the human brain.

Through multimodal data integration that combines PPI, whole-cell proteomic, and phosphoproteomic datasets in stem cell-derived neurons generated here, with the latest genomic and transcriptomic atlases in psychiatry, we identified a subset of neuron-specific PPI networks, or modules, that unify genetic, transcriptomic, proteomic, and/or drug response perturbations linked to SCZ. The neuropsychiatric relevance of these modules was validated through whole-cell proteomics, revealing they are down-regulated in neurons from 22q11.2 deletion carriers diagnosed with SCZ. This is consistent with the directionality of loss-of-function (LoF) coding mutations in the SCZ proteins anchoring the networks, as well as with the significant enrichment of LoF mutations throughout the networks. Furthermore, we demonstrated therapeutic relevance by showing that these modules are targets of GSK3 inhibition, as revealed via phosphoproteomics.

By meta-analyzing our data, we defined three distinct modules that exhibited the most convergent signals across the multimodal analyses, including networks centered on GRIA3, HCN4, and a highly overlapping set of networks anchored by SETD1A, TRIO, RB1CC1, AKAP11, and SRRM2. These modules include proteins that are traditionally linked to SCZ— such as those involved in glutamate receptor function [20] and voltage-gated cation channels [21]. Additionally, they include proteins with diverse biological functions, subcellular localizations, and genetic associations. Despite this functional diversity, these proteins converge on key cellular processes, including transcriptional and post-transcriptional regulation and the maintenance of cytoskeletal integrity.

Through multimodal data generation and integrative analyses, we present a foundational protein pathway model of SCZ and identify neuron-specific protein-protein interaction modules that concentrate SCZ-related perturbations and therapeutic responses. The model offers a robust framework for uncovering convergent mechanisms and drug targets in SCZ, which can also serve as a paradigm for analogous work in other neuropsychiatric disorders.

## Results

### Overview of modeling datasets

To establish a framework leveraging neuron-specific PPI for integrating genetic, transcriptional, and proteomic perturbations in SCZ, we generated multimodal datasets comprising single-cell transcriptomic and proteomic profiles from stem cell-derived excitatory and inhibitory neurons, as well as PPI data from cell models (comprising over four billion cells) and adult cortex tissue obtained from four donors (**Figure 1**). Rigorous experimental and computational quality control and harmonization of the PPI, including comparison against primary brain tissue data [22], identified neuron-specific sub-networks most likely to reflect biologically conserved functions in the adult brain. Building on this foundation, we developed an integrative model incorporating exome sequencing [3], genome-wide association studies (GWAS) [2], and single-nucleus RNA-seq (snRNA-seq) [7] datasets derived from individuals with SCZ, uncovering distinct SCZ modules with the highest convergence across multimodal analyses. The modules were subsequently validated using proteomic and phosphoproteomic data from neuronal models of the 22q11.2 CNV and through direct testing of known therapeutics, including GSK3 inhibitors.

**Figure 1.**
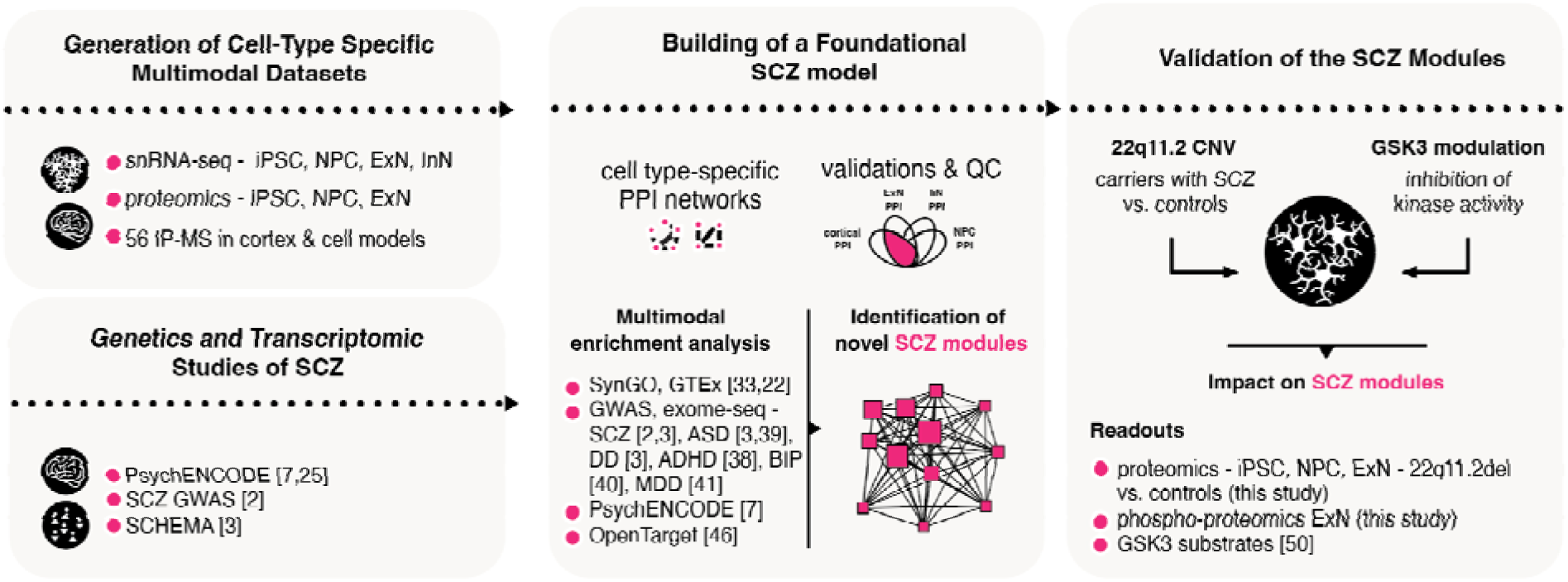
Study outline. Elements to building and validating a foundational neuronal protein network model unifying genetic, transcriptional, and proteomic perturbations in SCZ. snRNA-seq, single-nucleus RNA sequencing; iPSC, induced pluripotent stem cells; NPC, neural progenitor cells; ExN, excitatory neurons; InN, inhibitory interneurons; IP-MS, immunoprecipitation-mass spectrometry; SCZ, schizophrenia; GWAS, genome-wide association studies; SCHEMA, Schizophrenia Exome Sequencing Meta-Analysis; PPI, protein-protein interaction; QC, quality control; SynGO, Synaptic Gene Ontologies; GTEx, Genotype-Tissue Expression; ASD, autism spectrum disorder; DD, developmental delay; ADHD, attention-deficit/hyperactivity disorder; BIP, bipolar disorder; MDD, major depressive disorder; CNV, copy number variant.

### Establishing iPSC-derived NPC and ExN as models of the human cortex for multimodal analyses and interaction proteomics in SCZ

Analyses identifying genes with a significant burden of loss-of-function mutations in SCZ cases versus controls (e.g., SCHEMA study [3]) offer valuable protein-level anchors into risk-associated pathways, which are also enriched for common variants identified through GWAS [2]. Despite the value of proteomic datasets in psychiatry, they remain scarce due to challenges in scaling stem cell-based neuronal models, limited antibody availability, inherent variability in mass-spectrometry-based results, and the lack of systematic computational strategies and frameworks for harmonizing and integrating datasets.

To bridge this gap in the field, we established a protocol for scalably and systematically mapping PPI of proteins encoded by SCZ rare variant genes prioritized by SCHEMA. Our approach integrates two complementary and synergistic contexts: postmortem cortical homogenates from adult donors and stem cell-derived neuronal models (**Figure 2A**). This strategy addresses two key limitations in the field. First, by optimizing immunoprecipitation buffer conditions, we capture not only stoichiometric interactions but also dynamic and higher-order (second- and third-degree) interactions, enabling a more comprehensive mapping of pathway relationships. Second, by integrating primary tissue with cell-based models, we mitigate the limitations inherent to each system. While interactions identified in primary cortical tissue reflect molecular mechanisms present in the adult brain, they lack cell type specificity due to the cortex’s diverse cellular composition. In contrast, stem cell-based models, which approximate the developing human brain, offer greater cell type resolution for investigating protein interactions.

**Figure 2.**
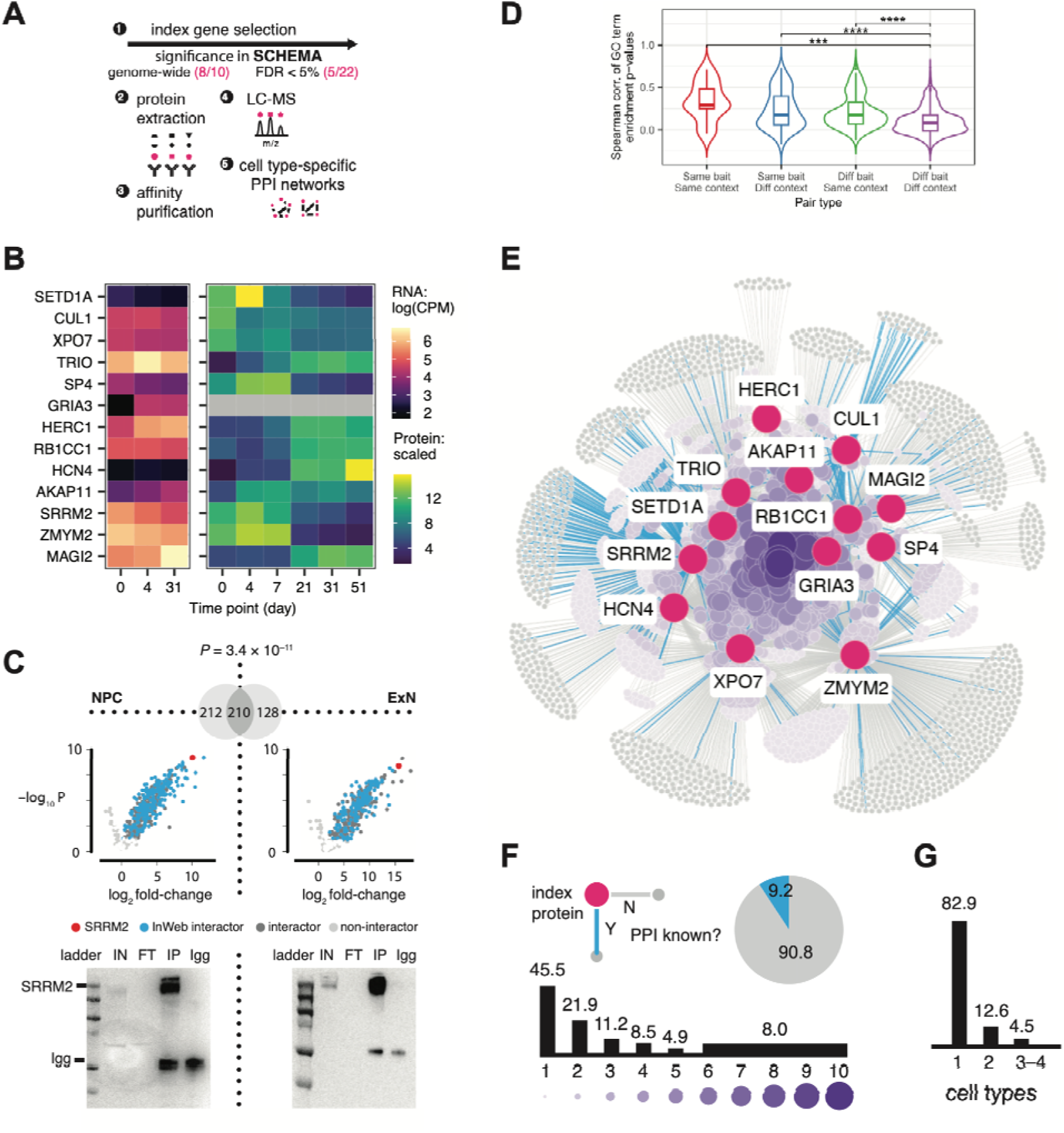
Identifying protein pathway relationships through protein-protein interactions of rare variant schizophrenia risk genes in *postmortem* cortex and stem cell-based neuronal models. **(A)** Workflow to generate PPI data across multiple cellular contexts. **(B)** RNA and protein expression heat maps for the SCZ index genes in iPSC-derived differentiating ExN. RNA expression was calculated by pseudo-bulking snRNA-seq data; protein expression was measured via whole-cell proteomic profiling (GRIA3 was not detected in this dataset). CPM, counts per million. **(C)** Volcano plots (top) and western blots (bottom) for two IP-MS experiments of SRRM2 in iPSC-derived NPC vs. ExN. Overlap of significant (FDR < 0.1) SRRM2 interactors detected in the two cell types are shown in the Venn diagram above the volcano plots. Expected molecular weights of SRRM2 and IgG chain are marked to the left of the western blot images; IN, input; FT, flow-through; IP, immunoprecipitation; Igg, IgG control. **(D)** Comparison of IP-MS dataset pairs stratified by index protein and cellular context. Spearman correlation between each pair was calculated based on enrichment p-values of GO Biological Process terms. *** and **** indicate FDR < 0.001 and 0.0001, respectively, calculated using two-tailed Wilcoxon rank-sum tests. **(E)** Unified PPI network from 56 IP-MS datasets across cortex, NPC, ExN, and InN. Nodes indicate index (red) or interactor proteins; edges indicate observed PPI. **(F)** Percentages representing novelty of the interactions assessed using PPI databases (top) and recurrence of the interactors across index proteins (bottom). **(G)** Percentages representing recurrence of the interactions across cell types (i.e., cellular contexts).

Given that cortical excitatory neurons account for the majority of SCZ risk within their expression profiles [7], we utilized a model of induced pluripotent stem cell (iPSC)-derived excitatory neurons (ExN) to study neuron-specific PPI of the rare variant SCZ risk genes. Specifically, we combined *NGN2* induction with small molecule patterning to generate reproducible and scalable populations of glutamatergic-like neurons, in quantities sufficient for MSLbased proteomic analyses [23, 24] (**Figure S1A**). As a comparison, we also included a distinct stem cell-based model of inhibitory interneurons (InN) [25], which are also implicated in SCZ, albeit to a lesser extent than ExN [7].

We evaluated the homogeneity and identity of these neuronal models using single-nucleus transcriptomics (**Table S1**). First, we analyzed snRNA-seq data of iPSC (day 0), proliferating neural progenitor cells (NPC; day 4), and fully differentiated ExN (day 31) to confirm that they formed distinct clusters expressing canonical pluripotency, progenitor, and excitatory neuronal markers, respectively. Notably, the ExN clustered separately from the InN, which expressed inhibitory markers (**Figure S1B**). We then applied ProjecTILs [26] to map the transcriptomic profiles of these cells onto lineages observed in a reference dataset of prenatal and postnatal human cortex [27]. As expected, the iPSC mapped to non-neuronal (vascular) cells in the cortex, while the NPC, ExN, and InN projected onto neuronal lineages. The ExN clustered more closely with excitatory lineages, while the InN mapped to inhibitory lineages (**Figure S1C**). These projections also revealed a developmental shift, with the NPC transitioned into more mature ExN resembling cortical excitatory neurons in mid to late prenatal stages (**Figure S1C,D**).

In parallel, we conducted TMT-labeled whole-cell proteomic profiling of differentiating ExN to quantify the expression of >8,000 proteins across six differentiation time points. Consistent with the snRNA-seq results, the iPSC (day 0), NPC (days 4-7), and ExN (days 21-51) samples expressed the expected cell type markers and were clearly separated according to their differentiation stages in principal component analysis (**Figure S1E,F**). We also performed k-means clustering to categorize all quantified proteins into five clusters based on their expression profiles throughout differentiation (**Figure S1G** and **Table S2**). Gene set enrichment analysis of the clusters with elevated expression in iPSC (“Pluripotency”), NPC (“Progenitor”), or ExN (“Neuronal”) revealed that they correspond to known functional transitions occurring during neurodevelopment, with the “Neuronal” cluster showing significant enrichment for neuron-specific processes (**Figure S1H** and **Table S3**).

Overall, both the transcriptomic and proteomic profiling data demonstrate that iPSC-derived NPC and ExN serve as reliable proxies for studying SCZ relevant pathway relationships of SCHEMA genes across chronological stages of brain development, with ExN approximating neurons in the perinatal cortex.

### Generating PPI data of rare variant SCZ risk genes in postmortem cortex and stem cell-based neuronal models

Before mapping the PPI of rare variant SCZ risk genes, we first confirmed the expression of their transcripts and encoded proteins in the human cortex and stem cell-based neuronal models. We then tested commercially available antibodies for their ability to immunoprecipitate the endogenous proteins in these cellular contexts. In total, we prioritized 13 proteins encoded by the top SCHEMA genes (eight exome-wide significant, five with false discovery rate [FDR] < 0.05) with robust expression and competent antibodies, including SETD1A, CUL1, XPO7, TRIO, SP4, GRIA3, HERC1, RB1CC1, HCN4, AKAP11, SRRM2, ZMYM2, and MAGI2 (hereafter referred to as index proteins; **Figure 2A,B**, **Figure S2A,B**). In line with known developmental trajectory of gene expression [28], some index proteins were more highly expressed in NPC (e.g., SETD1A, SP4, and SRRM2), while others were more abundant in mature ExN (e.g., GRIA3, HERC1, and HCN4). This suggests the index proteins may play functionally important roles at different stages of neurodevelopment, further motivating our investigation of their PPI in both NPC and ExN.

To generate pathway-focused PPI data, including second- and third-degree interactions in addition to stoichiometric ones for each index protein, we performed co-immunoprecipitation (co-IP) experiments using an antibody validated for immunoprecipitation in a modified Ripa buffer with 1% NP-40 or 0.5% n-Dodecyl β-D-maltoside (**Methods**). These experiments were conducted in adult tissue (*postmortem* cortical homogenates), and neuronal cell models (NPC, ExN, or InN), (**Figure 1A** and **Table S4**). All experiments were carried out in triplicates with matched controls (i.e., IP using non-specific IgG antibodies). Protein abundances in the IP samples were quantified by label-free liquid chromatography-tandem mass spectrometry (LC-MS/MS). We used Genoppi [24] to perform quality control (QC) and analyze the resulting IP-MS dataset from each experiment. Datasets in which the index protein was not enriched in the index protein IP compared to controls (log_2_ fold change [FC] > 0 and FDR < 0.1) were excluded from further analysis. In the remaining datasets, we defined all other enriched proteins (log_2_ FC > 0 and FDR < 0.1) to be the significant interactors of the index protein. We also cross-referenced the datasets with known interactors of the index proteins curated in PPI databases [16, 29–31] to assess the potential novelty of the observed PPI. In total, we analyzed 246 independent IP samples derived from four adult postmortem cortices and over 4 billion cells (using 2–5 mg of protein lysate per replicate) to generate 56 IP-MS datasets for the 13 index proteins, with each index protein having between 2-6 IP-MS datasets (i.e., 6-18 IP replicates) across the four cellular contexts (**Table S4**). **Figure 2C** highlights two of the datasets for SRRM2 in NPC and ExN as illustrative examples; all 56 datasets are provided in **Table S5**.

We systematically examined the QC metrics of the 56 IP-MS datasets to evaluate the robustness and comparability of the data across varying conditions. As expected, datasets generated at different sites (**Methods**) and in distinct cellular contexts showed variability in the number and pattern of detected proteins (**Figure S3A,B** and **Table S4**). However, after implementing a standardized Genoppi analysis workflow to filter the data and define the index protein interactors, these differences were mitigated and the QC metrics of the interactors became comparable across conditions (**Figure S3A,C**). To assess the biochemical reproducibility of the identified PPI, we performed western blot analysis for a subset of 137 and 118 interactions in NPC and ExN, respectively, to verify their persistence in independent IP experiments (**Table S6**). We successfully detected and replicated 64.3% (164/255) of the tested interactions across the two cell types. Reassuringly, the replication rates for previously reported vs. newly identified interactions were comparable (62.8% and 64.6%, respectively; **Figure S4A**). As a group, replicated interactions exhibited higher log_2_ FC and greater recurrence across the IP-MS datasets compared to those not validated by immunoblotting (**Figure S4B**). However, these trends were relatively subtle, as we were able to replicate many interactions with low log_2_ FC or detected in only a single IP-MS dataset, while certain interactions with high log_2_ FC and recurrence failed to replicate. Interactions that we could not replicate are likely due to a combination of factors such as the lower sensitivity of western blot compared to mass spectrometry, as well as the specificity and sensitivity of the antibodies used to detect the interactors. Furthermore, at the selected analysis cutoff (FDR < 0.1) for the IP-MS data, approximately 10% of the interactions are anticipated to be false positives. These likely arise from spurious in vitro interactions that would not replicate in subsequent WB analyses, further contributing to these observations.

To confirm that the identified PPI captured real protein pathway relationships despite other sources of biological and technical variability, we performed pairwise comparisons between the 56 IP-MS datasets. We stratified the dataset pairs by the two biological variables of interest, index protein and cellular context, to test whether datasets that shared these variables were more similar. Across several protein-level metrics, datasets for the same index protein in the same cellular context (same/same) generally showed the best agreement, followed by those for the same index protein in different contexts (same/different), different index proteins in the same context (different/same), and different index proteins in different contexts (different/different; **Figure S4C,D** and **Table S7**). The same trend persisted when we assessed pathway-level agreement between datasets based on their enrichment patterns across Gene Ontology (GO) terms [24, 32]: the same/same, same/different, different/same, and different/different dataset pairs showed decreasing correlations (median Spearman’s ρ = 0.293, 0.176, 0.173, and 0.0797, respectively), with the different/different pairs being significantly less correlated compared to the other three pair types (FDR < 1e-3, using two-tailed Wilcoxon tests; **Figure 2D** and **Table S8**). These results support the robustness of our IP-MS approach despite experimental variability, and indicate that both the index protein, and to a lesser extent cellular context, account for substantial variability in the data. Importantly, the GO term enrichment patterns validate that the PPI data effectively cluster the identified proteins with established biological pathways. Building on this and considering the extensive array of newly identified PPIs, we hypothesized that these networks likely encompass additional neuronal pathways that remain uncharacterized. In downstream analysis, we aimed to further distill the PPI data to identify and validate such pathways associated with SCZ.

After QC, we aggregated the PPI identified across all IP-MS datasets into a unified network, which includes 6,297 interactions involving 2,666 unique interactors of the 13 index proteins (**Figure 2E**; **Tables S9** and **S10**). Even though we replicated many (580) previously known interactions in PPI databases, over 90% of the interactions in the network had not been reported at the time of this study (**Figure 2F, top**). This aligns with previous findings from our group and others [17–19, 24], underscoring the limited number of cell type-specific PPI datasets in the current literature. Furthermore, the network shows a high level of connectivity, with over 50% of the interactors associated with multiple index proteins, suggesting that there may be functional convergence between them (**Figure 2F, bottom**).

When partitioning the PPI network by cellular contexts, 17.1% of the interactions were identified in more than one cell type, while 82.9% were identified in only one cell type (i.e., context-specific; **Figure 2G** and **Figure S5A**). Interactions identified across the stem cell-based models showed higher degree of overlap (Jaccard index = 0.12-0.25) compared to those identified in the cellular models vs. *postmortem* cortex (Jaccard index = 0.016-0.046). Similarly, most context-specific interactions were identified in the cortex rather than in the cellular models. These patterns are expected given the cell type diversity, complexity, and older developmental age of cortical tissue, as well as technical differences between the IP-MS experiments (**Methods**). Despite these differences, the overlaps between the cellular models and cortex were still highly significant (hypergeometric *P* < 2.9e-46; **Table S11**), and 352 interactions identified in the models were also found in the cortex, indicating that pathway-level insights gleaned from neuronal cell models can mirror those in complex tissues of the the human brain.

To quantitatively assess whether the sub-network derived from each cellular context captures biological processes in the developing and adult brain, we first conducted tissue enrichment analysis using GTEx gene expression data from adult donors [22]. We found that the Cortex and ExN networks were both highly enriched for tissue-specific genes in the central nervous system (**Figure S5B** and **Table S12**). By contrast, the NPC network, representing earlier stages of neurodevelopment, showed enrichment for tissues predominantly composed of non-neuronal cell types. When repeating the analysis using brain region-specific genes, only the Cortex and ExN networks were enriched for the genes specifically expressed in the frontal cortex (**Figure S5C** and **Table S13**). SynGO gene set analysis [33], which focused on synaptic genes and functions, revealed a similar pattern: while the Cortex and ExN networks both exhibited significant enrichment for a wide range of synaptic processes, the enrichment signals for NPC and InN networks were mostly concentrated in metabolic processes that activate earlier in synapse development (**Figure S5D** and **Table S14**).

Overall, these results suggest that PPI derived from *in vitro* models can reflect biological characteristics of different stages of neurodevelopment. Furthermore, the ExN network displays remarkably similar enrichment patterns compared to the Cortex network, indicating that it can also serve as a proxy to study SCZ-relevant neuronal pathways in the adult brain.

### The PPI network captures genetic risks of neuropsychiatric and neurodevelopmental disorders

To assess whether the PPI network anchored by the 13 index proteins captures genetic signals of SCZ, we first examined the network genes for any previously reported associations with SCZ from exome sequencing and GWAS. In total, 11 network genes have suggestive rare variant associations in SCHEMA [3], including five genes at FDR < 0.05 (*P* = 8.23e-5) and another six at FDR < 0.25 (*P* = 6.94e-4; **Figure S6A** and **Table S10**). For common variant associations, 20 network genes were prioritized by statistical fine-mapping and eQTL analyses from SCZ GWAS loci reported by the Psychiatric Genomics Consortium [2], with 227 additional genes falling within significant genomic regions in general (linkage disequilibrium [LD] r^2^ > 0.1 clumps ± 50kb; **Table S10**). Given the shared genetic signals between SCZ and neurodevelopmental disorders, we also referenced a recent exome sequencing study of autism spectrum disorder (ASD) and developmental delay (DD) [34] to annotate >200 network genes associated with these disorders at FDR < 0.05 (*P* = 5.54e-4 and 1.44e-3 for ASD and DD, respectively; **Table S10**).

Given that many genes in the PPI network were implicated in genetic studies of SCZ and ASD/DD, we next assessed whether the network is statistically enriched for genetic risk of these disorders compared to the protein-coding genome. We tested genes from the entire network (Union), the sub-networks derived from each cellular context (Cortex, NPC, ExN, InN), and the overlap sub-networks between each cellular model and cortex (NPC-Cortex, ExN-Cortex, InN-Cortex) for enrichment of rare coding variants linked to SCZ, ASD, and DD [3, 34]. While all tested networks showed significant enrichment for ASD and DD, only the ExN network was enriched for SCZ risk (FDR < 0.05, using one-tailed Kolmogorov-Smirnov test; **Figure 3A** and **Table S15**); the Union and InN networks also showed nominal enrichment (*P* < 0.05) for SCZ. These findings align with known genetic correlations between these disorders and their developmental trajectories: biological pathways associated with ASD/DD may influence early cortical development (e.g., the NPC network), whereas pathways linked to SCZ may be more relevant to later stages in more mature neurons (e.g., the ExN and InN networks). Furthermore, the lack of SCZ enrichment in the Cortex network suggests that the inclusion of non-neuronal cell types in cortical homogenates may have diluted the neuron-specific SCZ signal.

**Figure 3.**
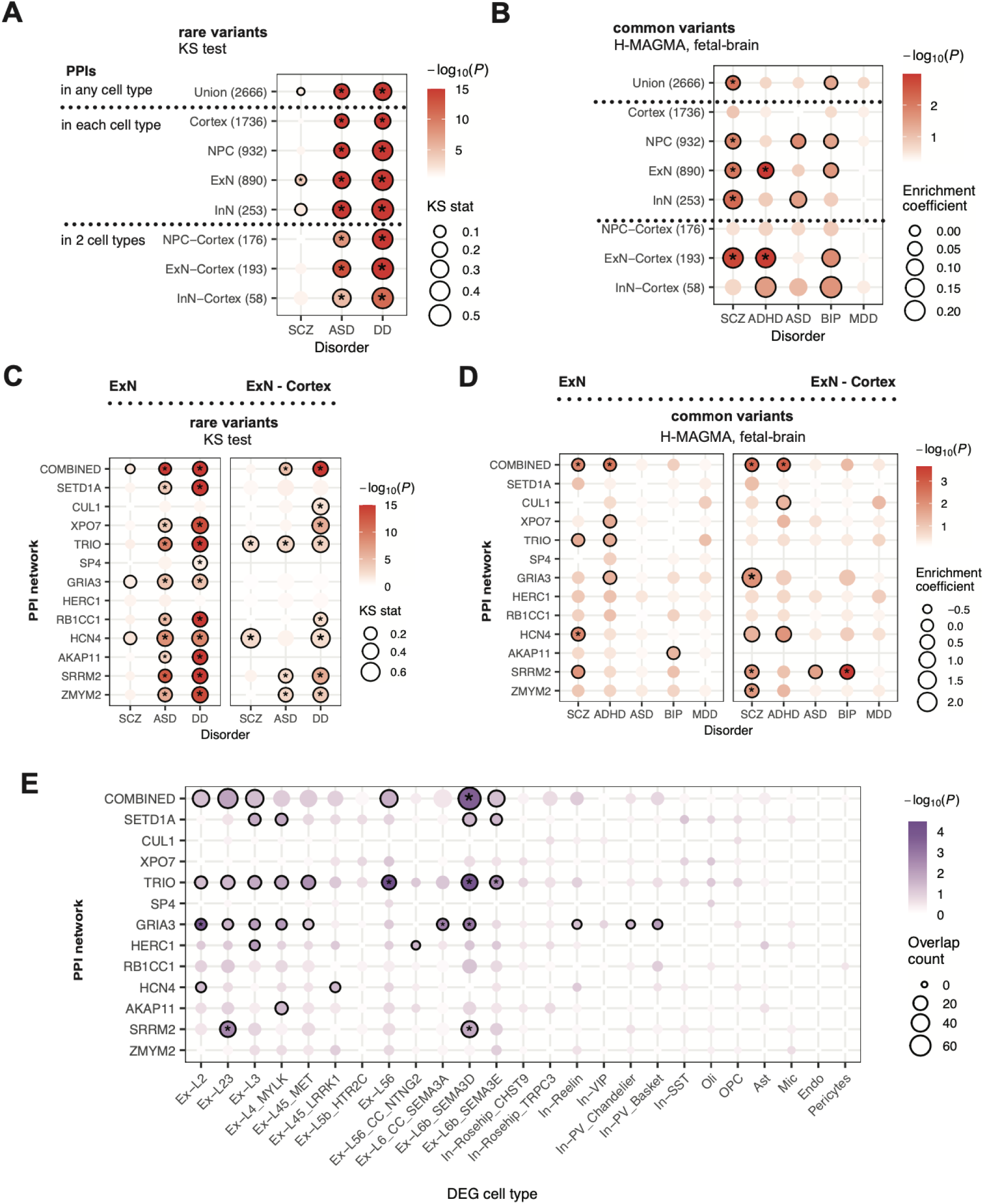
The ExN PPI sub-networks are enriched for genetic variants and transcriptional perturbations observed in SCZ individuals. **(A)-(B)** Rare and common variant enrichment of the combined PPI networks (across index proteins) compared against the protein-coding genome. **(C)-(D)** Rare and common variant enrichment of the ExN and ExN-Cortex sub-networks compared against the neuronal proteome. **(E)** Enrichment of cell type-specific SCZ DEGs in the ExN sub-networks compared against the neuronal proteome. Black border indicates *P* < 0.05, asterisk indicates FDR < 0.05.

Next, we used H-MAGMA [35, 36], Hi-C data from the fetal brain [37], and GWAS data of SCZ [2], attention-deficit/hyperactivity disorder (ADHD) [38], ASD [39], bipolar disorder (BIP) [40], and major depressive disorder (MDD) [41] to evaluate common variant enrichment of the networks. The Union and the cellular model-derived networks (NPC, ExN, InN) were all enriched for risk of SCZ compared to the protein-coding genome (FDR < 0.05; **Figure 3B** and **Table S16**). In contrast, the Cortex network did not show enrichment unless intersected with the ExN network (ExN-Cortex). In fact, the Cortex network showed no enrichment even when we repeated the H-MAGMA analysis using Hi-C data from the adult brain [42] (**Table S16**). Across other disorders, the ExN and ExN-Cortex networks were significantly (FDR < 0.05) linked to ADHD; several networks also showed nominal enrichment (*P* < 0.05) for ADHD, ASD, and/or BIP (**Figure 3B** and **Table S16**). Combined with the rare variant enrichment results, these findings reinforce that PPI networks derived from neuronal models can capture genetic signals of SCZ and other brain disorders. Importantly, the ExN network, which most closely resembles the Cortex network in GTEx and SynGO analyses (**Figure S5B-D**), shows consistent signal for SCZ in both rare and common variant analyses. By intersecting the ExN and Cortex networks, we were able to focus on cell type-specific PPIs in the excitatory neurons of the adult cortex, leading to a robust common variant enrichment for SCZ in the ExN-Cortex network.

### The ExN sub-networks are enriched for genetic variants and transcriptional perturbations observed in individuals with SCZ

Having established that both the ExN and ExN-Cortex networks are enriched for genetic risks of SCZ, we further examined whether these signals are broadly distributed across the networks or concentrated in certain sub-networks. We repeated the rare and common variant enrichment analyses on the combined (across index proteins) and index protein-specific sub-networks, comparing them to other genes expressed in the neuronal proteome (i.e., detected in the whole-cell proteomics dataset of differentiating ExN; **Figure S1E-H**). This conditional background allowed us to more conservatively assess network enrichment after accounting for the neuronal context of the ExN model.

In the rare variant analysis, three ExN networks (combined, GRIA3, HCN4) were nominally enriched (*P* < 0.05) for SCZ, while two ExN-Cortex networks (TRIO, HCN4) showed significant enrichment (FDR < 0.05; **Figure 3C** and **Table S17**). Comparatively, the ExN networks were generally more enriched for ASD and DD compared to their ExN-Cortex counterparts, which could be due to better representation of perinatal interactions or better statistical power (due to larger network sizes) in the ExN networks. In the common variant analysis, two ExN networks (combined, HCN4) and four ExN-Cortex networks (combined, GRIA3, SRRM2, ZMYM2) were enriched (FDR < 0.05) for SCZ risk, with additional networks (ExN: TRIO, SRRM2; ExN-Cortex: HCN4) reaching nominal significance (*P* < 0.05; **Figure 3D** and **Table S18**). Both the ExN and ExN-Cortex combined networks were also significantly enriched for ADHD, and the ExN-Cortex SRRM2 network was also enriched for BIP. These findings suggest that the networks may be capturing convergent pathways both between SCZ and other brain disorders, and between rare and common variants linked to SCZ. In particular, the combined network along with the TRIO, GRIA3, and HCN4 networks all displayed recurrent SCZ signals across the rare and common variant analyses.

To determine whether the PPI networks also concentrated transcriptional alterations linked to SCZ, we used data from a PsychENCODE snRNA-seq study that identified differentially expressed genes (DEG) in SCZ cases vs. controls across 25 annotated cell types in *postmortem* cortex [7]. Several ExN networks (combined, TRIO, GRIA3, SRRM2) were found to be enriched for DEG in both superficial and deep layer cortical excitatory neurons (FDR < 0.05, using hypergeometric test and the neuronal proteome background; **Figure 3E** and **Table S19**). The enrichment signals in the combined, TRIO, and GRIA3 networks were predominantly driven by down-regulated genes in SCZ individuals compared to controls, which was consistent with the broader trend of transcriptome-wide down-regulation observed in the snRNA-seq study and the directionality of loss-of-function mutations that prioritized the SCZ index proteins anchoring the networks (**Figure S6B**). In contrast, the SRRM2 network showed greater enrichment for up-regulated genes in SCZ, a finding that was also replicated in the ExN-Cortex SRRM2 network (**Figure S6C**). Overall, many of the index protein-specific sub-networks enriched for SCZ-associated genetic variants also showed enrichment for transcriptional dysregulation observed in the brains of SCZ individuals. We hypothesized that these sub-networks may represent functional modules that include previously unrecognized SCZ-associated genes and pathways, and that perturbations in these modules could provide critical insights into the molecular mechanisms underlying the disorder.

### Model validation: the ExN sub-networks are enriched for proteomic perturbations in iPSC-derived neurons from 22q11.2del carriers with SCZ

To validate that the ExN sub-networks represent functional modules dysregulated in SCZ, we tested whether these networks also concentrate SCZ-associated perturbations in neurons derived from individuals with genetically well-defined SCZ. We focused on perturbations driven by the chromosome 22q11.2 deletion (22q11.2del), a penetrant CNV linked to SCZ, as it is independent of the rare variants used to prioritize the SCZ index proteins for this study [3, 5, 43] and has one of the highest effect sizes observed in SCZ. We selected iPSC lines from eight previously characterized donors, including four 22q11.2del heterozygous carriers diagnosed with SCZ and four age- and sex-matched controls, and differentiated them into NPC and ExN [44]. TMT-labeled proteomic profiling was then performed to compare protein expression in iPSC, NPC, and ExN containing the 22q11.2del vs. controls (**Figure 4A**).

**Figure 4.**
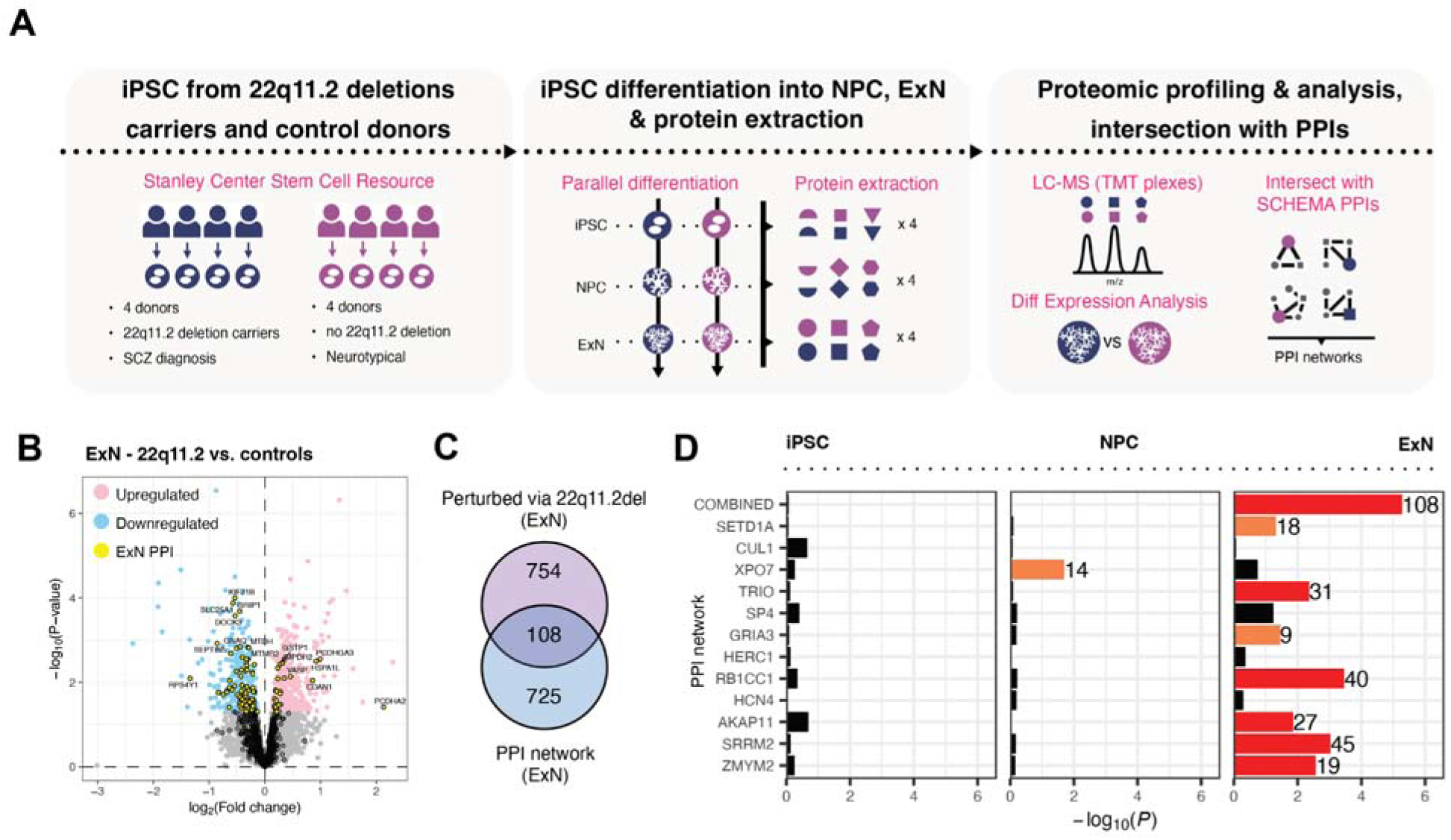
Proteomic profiling of iPSC-derived neurons from 22q11.2del carriers with SCZ revealed dysregulation in the ExN PPI sub-networks. **(A)** Workflow to generate proteomic data for studying the 22q11.2del. **(B)** Volcano plot showing differential expression of proteins in 22q11.2del vs. control ExN, overlaid with the ExN PPI network. DEP (*P* < 0.05) that were up- or down-regulated in 22q11.2del ExN are shown in pink or blue, respectively. **(C)** Overlap between the DEP in 22q11.2del ExN vs. the ExN PPI network. **(D)** Overlap enrichment between the DEP in 22q11.2del iPSC, NPC, or ExN (column facets) vs. the ExN PPI sub-networks. Red bars indicate FDR < 0.05; orange bars indicate *P* < 0.05; number of overlapping proteins is shown to the right of each colored bar.

The proteomic data revealed that the 22q11.2del did not affect overall differentiation efficiency or timing: all samples clustered according to cell state in principal component analysis and expressed the appropriate cell type markers, regardless of deletion status (**Figure S7A,B**). As a positive control, we confirmed that proteins encoded by genes in the 22q11.2del region exhibited lower expression in the deletion samples compared to the controls (*P* = 1.1e-8, using two-tailed Wilcoxon rank-sum test; **Figure S7C**), indicating that expression differences for gene sets of interest could be detected in this dataset. We calculated differential expression statistics for all proteins in the 22q11.2del vs. control iPSC, NPC, or ExN (**Figure S7D** and **Table S20**), and used these statistics in gene set enrichment analysis to identify biological processes perturbed by the deletion (**Figure S7E** and **Tables S21**). Proteins associated with translation and the cytosolic ribosome were most affected by the deletion in iPSC and NPC, but in opposite directions (up-regulated in iPSC and down-regulated in NPC). In ExN, proteins related to nervous system development, synaptic transmission, axonogenesis, and mitochondrial OXPHOS were down-regulated in ExN carrying the deletion. These findings align with the functional specialization of differentiating neurons and may reflect the complex clinical manifestations of the 22q11.2del syndrome, which impacts both psychiatric and broader developmental processes [45].

To link the proteomic changes caused by the 22q11.2del back to the SCZ signals in the ExN PPI networks, we assessed whether proteins in the networks were more likely to be perturbed by the 22q11.2del compared to other proteins detected in the experiment. First, we observed an enriched overlap between the ExN combined network and the differentially expressed proteins (DEP; nominal *P* < 0.05) in the 22q11.2del vs. control ExN (hypergeometric *P* = 5.3e-6; **Figure 4B,C** and **Table S22**). This enrichment was primarily driven by the down-regulated proteins in the 22q11.2del ExN (**Figure S6F**). In contrast, no significant overlap was found between the ExN network and DEP in iPSC or NPC (**Figure 4D**). Next, we analyzed the ExN index protein-specific sub-networks and observed similar patterns: five networks (TRIO, HERC1, AKAP11, SRRM2, ZMYM2) showed enriched overlap with all DEP in 22q11.2del ExN (FDR < 0.05; **Figure 4D**), while five additional networks (SETD1A, XPO7, SP4, GRIA3, RB1CC1) reached significance when considering only the down-regulated DEP in ExN (**Figure S7F**). Conversely, only the XPO7 network was enriched for down-regulated DEP in NPC and no network was enriched for DEP in iPSC. Finally, the ExN-Cortex combined, RB1CC1, and SRRM2 networks were also enriched for down-regulated DEP in 22q11.2del ExN (**Figure S6F**).

These results further support the notion that the ExN PPI network captures biological pathways altered in the brains of SCZ individuals. By integrating the network with multiple orthogonal datasets, we show that dysregulation of these pathways occurs at the genomic, transcriptomic, and proteomic levels across diverse genetic backgrounds. Furthermore, several ExN sub-networks (e.g., TRIO, GRIA3, HCN4, SRRM2) were consistently prioritized across the different analyses, suggesting that they may represent “SCZ modules” that contribute to convergent molecular machinery broadly dysregulated in SCZ, which could serve as potential targets for therapeutic interventions.

### Protein modules are convergent hubs for known therapeutic targets

To explore the hypothesis that the SCZ modules could serve as convergent hubs for therapeutic interventions, we first searched data from the Open Targets Platform [46] to identify 290 proteins in our PPI network that are known drug targets (**Table S10**). Among these proteins is GSK3B, a well-known lithium target whose inhibition has been shown to effectively manage symptoms in individuals with psychosis across diverse genetic backgrounds [47–49]. In both NPC and ExN, GSK3B interacts with TRIO and SRRM2, whose ExN sub-networks show SCZ signals across multiple analyses. Consistent with the PPI data, GSK3B was highly expressed in ExN compared to its paralog, GSK3A, which was not detected as an interactor of any index proteins (**Figure S8A**). Intrigued by these connections, we mapped known substrates of GSK3B [50] onto the ExN combined network and observed an enriched overlap (hypergeometric *P* = 1.0e-14; **Figure 5A, top**). Additionally, we performed an IP-MS experiment of GSK3B in ExN to identify additional GSK3B interactors (**Figure S8B** and **Table S23**), revealing an even more significant overlap between the ExN network and the interactors (*P* = 7.7e-160; **Figure 5A, bottom**).

**Figure 5.**
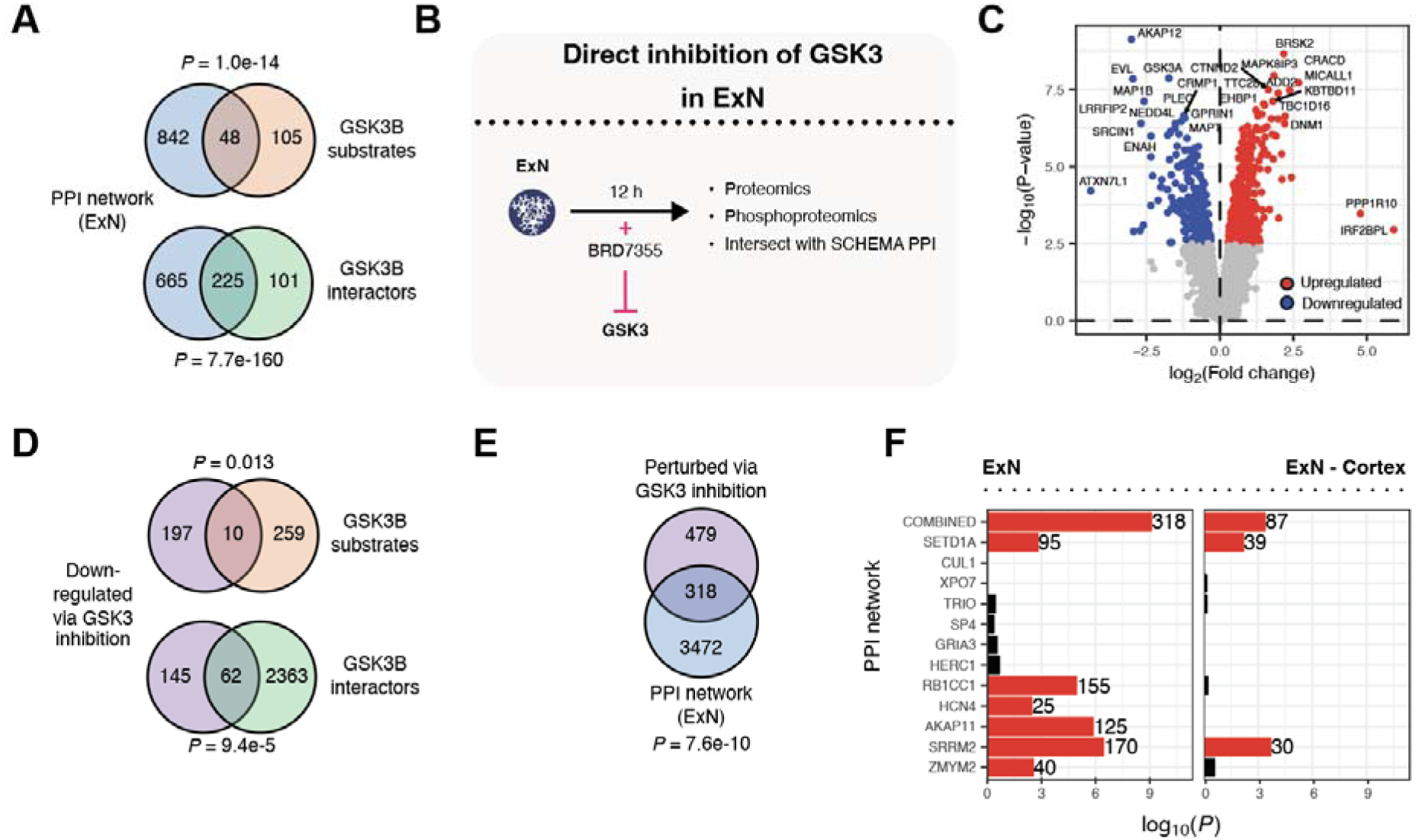
GSK3-mediated phosphorylation offers therapeutic opportunities to target SCZ modules contained within the ExN PPI sub-networks. **(A)** Overlap between the ExN PPI network vs. known GSK3B substrates or IP-MS-derived GSK3B interactors in ExN. **(B)** Workflow to study effects of GSK3 inhibition in ExN. **(C)** Volcano plot showing differential expression of phosphopeptides in GSK3 inhibitor-treated vs. control ExN. **(D)** Overlap between down-regulated phosphopeptides vs. GSK3B substrates or interactors. **(E)** Overlap between DE phosphopeptides vs. the ExN PPI network. **(F)** Overlap enrichment between the DE phosphopeptides vs. the ExN and ExN-Cortex PPI sub-networks. Red bars indicate FDR < 0.05; number of overlapping phosphopeptides is shown to the right of each red bar.

Considering these findings, we investigated whether proteins within the SCZ modules respond to GSK3B modulation in neurons. Due to the low potency/specificity ratio of GSK3B-specific inhibitors, and considering that GSK3A is expressed at a lower level in ExN compared to GSK3B, we treated ExN cells with a dual GSK3A/B inhibitor (BRD7355) and assessed alterations in both the proteome and phosphoproteome (**Figure 5B**). Specifically, we exposed ExN to a dose of BRD7355 corresponding to its IC50 for 12 hours, alongside a control treatment with the inactive enantiomer of BRD7355, and then performed TMT-labeled proteomic and phosphoproteomic profiling on the inhibitor-treated vs. control cells.

The proteomic data confirmed that the inhibitor-treated cells did not undergo apoptosis nor exhibit overexpression of stress-related proteins. Differential expression analysis of the inhibitor-treated vs. control samples revealed no significant changes in protein abundance at FDR < 0.05, indicating that BRD7355 had minimal impact on protein abundance in ExN (**Figure S8C**). In contrast, when we analyzed the matching phosphoproteome profiles, we identified 798 differentially expressed phosphopeptides (DE phosphopeptides; FDR < 0.05) from 461 unique genes in the inhibitor-treated vs. control samples (**Figure 5C** and **Table S24**). Reassuringly, the down-regulated phosphopeptides in inhibitor-treated cells significantly overlap with known GSK3B substrates and interactors (hypergeometric *P* = 0.013 and 9.4e-5, respectively; **Figure 5D**), while the up-regulated phosphopeptides did not (**Figure S8D**). This suggests that BRD7355 effectively inhibited the kinase activity of GSK3B in ExN, leading to a reduction in the phosphorylation of its substrates.

We also observed a significant overlap between the DE phosphopeptides and the ExN combined network (hypergeometric *P* = 7.6e-10; **Figure 5E**). Among the index protein-specific sub-networks, six showed enrichment for the DE phosphopeptides (SETD1A, RB1CC1, HCN4, AKAP11, SRRM2, ZMYM2; FDR < 0.05; **Figure 5F** and **Table S25**). The enrichment in the combined, SETD1A, and SRRM2 networks was also replicated in the analogous ExN-Cortex networks (**Figure 5F**). Distinct from the results for the direct substrates of GSK3B, the signals in the PPI networks were often explained by both up- and down-regulated phosphopeptides upon GSK3 inhibition (**Figure S8E**). This suggests that the networks were enriched not only for GSK3B substrates, but also for proteins more broadly downstream of the GSK3 signaling pathway. Furthermore, several sub-networks central to the SCZ modules (**Figure 6A,B**) were particularly susceptible to inhibition of GSK3-mediated phosphorylation in ExN, highlighting therapeutic opportunities to target genes within these modules. This approach could help narrow the broad range of lithium’s targets, which has shown variable efficacy in alleviating SCZ symptoms [51].

**Figure 6.**
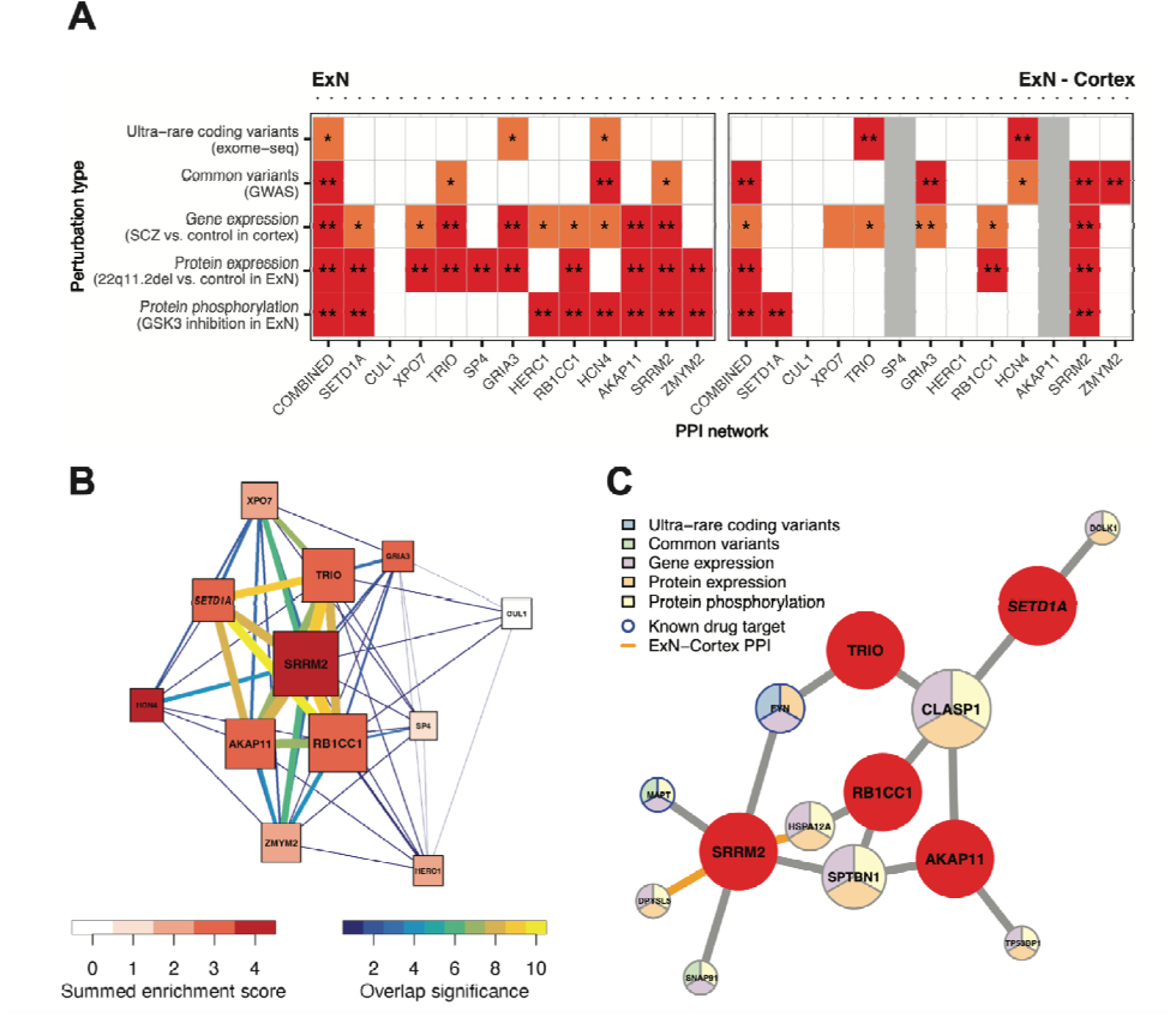
Multimodal PPI network enrichment analyses reveal distinct SCZ modules. **(A)** Enrichment of the ExN and ExN-Cortex sub-networks across five different types of SCZ-linked perturbations. Orange cell with single asterisk indicates the network reached nominal significance (*P* < 0.05) in the analysis; red cell with double asterisks indicates the network was significant at FDR < 0.05. **(B)** Convergence among the ExN sub-networks across orthogonal enrichment analyses. Summed enrichment score indicates the number of analyses in which the network reached at least nominal significance. Overlap significance between sub-networks indicates scaled −log_10_(*P*) calculated using hypergeometric tests; only significant overlaps (FDR < 0.05) were plotted. **(C)** Top convergent interactors (i.e., perturbed in 3 out of 5 datasets) in Module 3. Index proteins are shown in red; interactor proteins are color-coded based on SCZ-linked perturbation types and their sizes scale with the number of index proteins they interact with. Interactors that are known drug targets are highlighted by blue border; interactions that were identified in both ExN and cortex are highlighted in orange.

### Genetic, transcriptomic, and proteomic perturbations of SCZ converge on three distinct “SCZ modules” in ExN

To formally characterize the SCZ modules captured by the the ExN PPI sub-networks, we performed a meta-analysis to summarize their enrichment signals across five different types of SCZ-related perturbations, including ultra-rare coding variants from exome sequencing (protein-truncating or damaging missense variants; **Figure 3C**), common variants from GWAS (**Figure 3D**), gene expression changes in postmortem cortex (**Figure 3E**), protein expression changes in 22q11.2del neurons (**Figure 4D**), and protein phosphorylation changes in GSK3-inhibited neurons (**Figure 5F**). Seven of the index protein-specific ExN sub-networks (SETD1A, TRIO, GRIA3, RB1CC1, HCN4, AKAP11, SRRM2) emerged as the most convergent networks that were enriched for at least three perturbation types (**Figure 6A,B**). Some enrichment signals for these networks were also recapitulated in the analogous ExN-Cotex sub-networks, with the SRRM2 networks showing the most consistent agreement.

To assess whether these seven networks represent distinct or overlapping biological pathways, we first clustered them based on the degree of overlap between their network genes (**Figure 6B** and **Table S26**). Except for the overlap between the GRIA3 and HCN4 networks (FDR = 0.38, using hypergeometric test), all other overlaps between the seven networks were significant (FDR < 0.0072). The GRIA3 and HCN4 networks were also more distinct from the other five networks (FDR = 1.6e-28 to 0.0072 between HCN4 or GRIA3 vs. other networks; FDR = 7.1e-79 to 1.4e-56 between the other networks). Based on these findings, we defined three distinct SCZ modules: one centered on GRIA3 (Module 1), another on HCN4 (Module 2), and a third hub involving SETD1A, TRIO, RB1CC1, AKAP11, and SRRM2 (Module 3).

Consistent with this data-driven classification, REACTOME [52] pathway enrichment analysis revealed a similar clustering pattern, where the top enriched pathways differ between the three modules (**Figure S9A** and **Table S27**). Furthermore, filtering the newly defined modules for convergent interactors perturbed in at least two analyses prioritized a subset of 80 proteins with diverse perturbation profiles, nine of which are known drug targets (**Figure S9B** and **Table S10**). While Module 1 and Module 2 likely encompass proteins associated with glutamate receptors and voltage-gated cation channels—both known to play pivotal roles in SCZ [20, 21]—Module 3 stands out as a particularly intriguing regulatory hub. This module aggregates PPI of index proteins involved in diverse cellular processes, including splicing regulation, chromatin modification, and guanine nucleotide exchange [53–57]. Many interactions for the convergent interactors in Module 3 were replicated in both ExN and cortex; two of the index proteins, SRRM2 and TRIO, also interact with each other in both ExN and cortex (**Figure S9B**). These findings further implicate the functional relevance of this module in the human brain.

We further prioritized the top convergent interactors in Module 3, which contains nine proteins that are individually perturbed in three out of five analyses and are broadly implicated in neuronal function and integrity (**Figure 6C** and **Table S10**). These include two well-characterized drug targets: MAPT, a microtubule-associated protein harboring numerous single nucleotide polymorphisms (SNPs) linked to various neurodegenerative diseases [58]; and FYN [59], a gene associated with schizophrenia [60, 61] that encodes a kinase implicated in NMDA receptor regulation [59, 62]. Other convergent interactors identified in Module 3 include DCLK1, SPTBN1, DPSYL5, CLASP1, TP53BP1, CRMP, and HSPA12A. Although these genes encode proteins harboring loss-of-function mutations that have each been individually linked to neuronal disorders and schizophrenia through complementary mechanisms—such as neurite destabilization, defective axonal trafficking, or mislocalization of downstream signaling proteins [63–69]—they have not previously been recognized as a cohesive functional module.

In conclusion, we believe these newly identified SCZ modules capture distinct yet converging biological pathways that warrant prioritization in future therapeutic strategies. Notably, Module 3 highlights cytoskeletal integrity and regulation as a key mechanism in SCZ, while also presenting novel candidates for targeted intervention.

## Discussion

In this study we present a multimodal model that integrates protein-protein interactions with orthogonal data types, including genomics, transcriptomics, proteomics, and drug response data, to explore the genetic mechanisms of SCZ. This approach pinpoints a refined subset of genes and modules with strong cell type-specific connections to SCZ risk, while also revealing shared pathways with other neurodevelopmental disorders.

To build our model, we first mapped PPI for 13 rare variant SCZ index genes across complementary human brain cellular contexts—including postmortem cortical tissue, iPSC-derived NPC and ExN, as well as embryonic stem cell-derived InN. This allowed us to preserve the cell type specificity of interactions mapped in the cellular models while identifying shared biochemical pathways between the cellular models and adult brain tissue. We compared the PPI identified across independent IP-MS experiments to assess the well-documented variability in protein detection between both biological and technical replicates inherent to MS-based proteomics [70]. Reassuringly, the PPI identified for the same index protein and/or cellular context showed significantly higher correlation at both the gene and pathway levels compared to PPI identified for different index proteins and contexts, validating that these data capture true biological signals and pathways despite the well known caveats of cell models and proteomic analyses. By integrating 56 IP-MS datasets across the four cellular contexts - including four adult postmortem cortices and over 4 billion cells (using 2–5 mg of protein lysate per replicate) - we created an inclusive PPI network comprising 6,297 interactions involving 2,666 unique interactors, with over 50% connected to multiple SCZ index genes. Notably, over 90% of these interactions were previously unreported, indicating that the networks encompass additional neuronal pathways linked to SCZ that remain to be characterized, which is the focus of all subsequent analyses.

In GTEx tissue and SynGO enrichment analyses, the PPI sub-networks derived from cortex and ExN showed similar enrichment patterns for brain and synapse-specific genes and functions. In contrast, the NPC sub-network was enriched for non-neuronal genes and processes that activate earlier in cortical development. This suggests that the NPC and ExN networks reflect successive stages of neuronal development and that the ExN network can serve as a valuable proxy to study protein relationships in the adult brain despite the limited maturity of the in vitro model.

Consistent with these findings, testing the PPI networks for enrichment of common and rare variant risk genes associated with various neurodevelopmental and neuropsychiatric disorders revealed a striking distinction between the NPC and ExN networks. The ExN network was enriched for both rare and common variants linked to SCZ, while the NPC network was only enriched for the common variants. This emphasizes the pivotal role of excitatory neurons, especially those in deep cortical layers, in SCZ pathophysiology [7]. In contrast, rare variant genes linked to ASD and DD were similarly enriched in both the NPC and ExN networks. This distinction highlights how SCZ genetic risk is modulated across developmental stages and aligns with prior evidence of functional overlaps between SCZ and other neurodevelopmental disorders during critical neurodevelopmental windows [34]. These findings underscore the need to differentiate between general mechanisms that affect brain development and those specifically tied to SCZ in the adult brain.

Another intriguing finding is the absence of SCZ genetic signals in the Cortex network, especially compared to the networks generated from cellular models. As previous literature has established the relevance of the adult cortex in SCZ pathophysiology, we hypothesized that the absence of genetic enrichment could be explained by the lack of cell type specificity in the network. Indeed, when we intersected the PPI identified in cortical tissue with those found in the cellular models, the ExN-Cortex network emerged as one of the top networks in terms of common variant enrichment for SCZ. In other words, we show that the ExN-specific PPI network effectively preserves SCZ-relevant signals present in the primary tissue, while concurrently minimizing non-specific interactions.

Dissection of the ExN PPI network identified index protein-specific sub-networks that are enriched for rare or common variants linked to SCZ or are transcriptionally dysregulated in the postmortem cortical neurons of individuals with SCZ. Several sub-networks, including TRIO, GRIA3, HCN4, and SRRM2, show recurrent enrichment signals across these analyses, suggesting that they contain functional modules with unrecognized SCZ-associated genes. Interestingly, the directionality of transcriptional dysregulation can be network-specific: the TRIO and GRIA3 sub-networks are enriched for down-regulated genes in SCZ, while the SRRM2 sub-network is enriched for up-regulated genes.

To further validate the functional connections between the ExN sub-networks and SCZ, we examined how they are impacted by the 22q11.2 deletion, the most penetrant CNV linked to SCZ, as well as their response to inhibition of Glycogen Synthase Kinase-3 Beta (GSK3B), a kinase implicated in lithium-responsive signaling in SCZ [51]. Our analyses identified many sub-networks that are enriched for dysregulated proteins linked to 22q11.2del [5, 45, 71, 72]. Several of the same sub-networks are also enriched for phosphosites modulated by GSK3B inhibition in neurons.

By focusing on the ExN sub-networks that are most frequently perturbed across the SCZ-linked genetic, transcriptomic, proteomic, and phosphoproteomic datasets we analyzed, we identified three key SCZ modules: one centered on the GRIA3 network, another on the HCN4 network, and a third encompassing the SETD1A, TRIO, RB1CC1, AKAP11, and SRRM2 networks. These modules capture both established and novel biological pathways linked to SCZ, with Module 3 emerging as a regulatory hub with broad implications for neuronal integrity. Further investigation into the most perturbed genes in Module 3 could elucidate their roles in SCZ and their potential as therapeutic targets.

In summary, this study presents a novel framework that integrates diverse datasets with neuron-specific interaction proteomics to uncover convergent molecular mechanisms driving SCZ. By integrating proteomic and transcriptomic datasets from stem cell-derived neurons, postmortem samples, and drug response studies, we identified three neuron-specific "SCZ modules" that connect genetic risk to functional dysregulation across multiple modalities. These modules exhibit alterations in SCZ, including in the postmortem brains of individuals with idiopathic SCZ and in stem cell-derived neurons from carriers of the 22q11.2 CNV. Notably, the modules converge on pathways responsive to GSK3 inhibition, a kinase that can be targeted by small molecules, providing a mechanistic blueprint for therapeutic strategies aimed at restoring gene expression within these SCZ-associated modules. Our findings establish a paradigm for future efforts to functionally characterize SCZ-associated pathways and explore their therapeutic potential, offering broadly applicable strategies to address the diverse genetic architecture of SCZ and benefit individuals across the spectrum of genetic risk.

### Study limitations

This study has several limitations tied to both its design and the experimental methods employed. One major constraint was our choice to use specific antibodies for IP-MS of the index proteins rather than a tagging and overexpression approach, which would have required selecting specific isoforms and could risk non-endogenous expression. This decision restricted us to available antibodies that were compatible with IP, which could be circumvented in future research by using endogenous protein tagging. Another limitation arose from our method of comparing protein levels in target IPs against matched negative (IgG) controls to account for non-specific binding. An alternative approach, such as performing IPs in isogenic iPSC lines where target proteins are knocked out, could provide a more refined control but would also introduce challenges in maintaining cell viability and differentiation. Additionally, our IPs were conducted under conditions that captured both stable and transient protein interactions, yet our analysis does not distinguish between these types. Moreover, the mass spectrometry (MS) analysis we employed comes with known biases, such as incomplete protein detection and the underrepresentation of low-abundance or hydrophobic proteins, particularly those found in membranes. Finally, our PPI network is centered on 13 target proteins derived from three cellular models and postmortem samples, which limits its scope in capturing the full interactome of all relevant SCZ genes across diverse brain cell types. While our findings prioritize three SCZ protein modules, future studies will need to expand upon this work as more comprehensive genetic data and advanced cell modeling techniques become available. This will be essential for constructing a more complete, cell-type-specific PPI map that elucidates the functional interactions and pathways that are important for SCZ.

## Acknowledgements

We would like to thank Kris Dickson for fruitful discussions and Steve Hyman for helping to conceptualize this study and critically reading the manuscript. We thank the Harvard Center for Mass Spectrometry, the Taplin Mass Spectrometry Facility, and the Thermo Fisher Center for Multiplexed Proteomics, for discussions and generation of MS data. This work was supported by grants from the Stanley Center for Psychiatric Research, the US National Institute of Mental Health (R01 MH109903 and U01 MH121499), the Simons Foundation Autism Research Initiative (awards 515064 and 735604), the Lundbeck Foundation (R223-2016-721 and R350-2020-963), and the Novo Nordisk Foundation (NNF21SA0072102 and NNF21CC0073729). G.P. is supported by Simons Foundation Autism Research Initiative Bridge to Independence Award 00002804.

## Author Contributions

G.P., J.C.B, D.M., E.C., H.G. and U.D. carried out tissue culture. G.P., J.C.B, D.M., T.B., M.M., and D.R. tested antibodies and ran immunoblots. G.P., M.C., and K.W.L. executed IP experiments. K.W.L. and M.C. ran MS experiments and analysis at CNCR. M.W. provided and characterized GSK3 inhibitors. X.A. and N.H. performed snRNA-seq experiments. G.P. performed the rest of the experiments. Y.-H.H.H., P.P., and J.K.T.C. analyzed IP-MS data. Y.-H.H.H. and S.S. analyzed snRNA-seq data. Y.-H.H.H., P.P., and O.P. analyzed proteomic profiling data. Y.-H.H.H. performed the rest of the analyses. A.B.S., A.K., J.Z.L., R.N., T.W., and K.L. supervised and managed the study. N.F. raised funds to support this study and facilitated scientific collaborations. D.H. provided administrative support. K.L. designed and led the study. G.P. and Y.-H.H.H. wrote the manuscript with key contributions from KL and input from the co-authors.

## Competing interests

K.L. is equity holder at Sidera and consultant at Sidera and ZS Associates. A.K. is the owner of Kirkeby Cell Therapy APS, which holds royalty contracts on several patents related to stem cell differentiation and performs paid consultancy for companies working on pluripotent stem cell therapies.

**<u>Supplementary figures here</u>**

**<u>Supplementary tables here</u>**

## Methods

### Cell lines

NPC and ExN used in this study were differentiated from a human male iPSC line, iPS hDFn 83/22 iNgn2#9 (iPS3) [23, 24], with the exception of the 22q11.2del proteomic profiling experiment, which employed eight iPSC lines previously described and fully characterized in [44]: SCBB-1473 (control, male), SCBB-651 (deletion carrier, male), SCBB-798 (control, male), SCBB-1825 (deletion carrier, male), SCBB-1827 (control, female), SCBB-1652 (deletion carrier, female), SCBB-1648 (deletion carrier, female), and SCBB-1490 (deletion carrier, female). InN were differentiated from a human embryonic stem cell (ESC) line, RC17 (WA09 in [73]).

### Cell culture

All iPSC and ESC lines were seeded on a plate coated with GelTrex (LifeTechnologies, A1413301) adhesion matrix (1:100 in DMEM/F:12, Life Technologies, Inc., 11320033) at a density of 35,000 cells cm^-2^ in Stemflex media (Life Technologies, Inc., A3349401). Once the cells achieved 60% confluency, the monolayer was detached using Accutase (Life Technologies, Inc., A11105) and transferred onto new plates for cell expansion. Cells were incubated at 37°C and 5% CO_2_.

### Differentiation of iPSC to NPC and ExN

Human iPSC were differentiated to NPC and ExN by conditional expression of the neuralizing transcription factor *NGN2* combined with small molecule-mediated developmental patterning as previously described [23]. iPSC were plated at a density of 35,000 cells cm^-2^ with rock inhibitor Y27632 (Stemgent, 04-0012). Day 1 cells were differentiated in N2 media (Life Technologies, Inc.) supplemented with 10 μM SB431542 (Tocris, 1614), 2 μM XAV939 (Stemgent, 04-00046), and 100 nM LDN-193189 (Stemgent, 04-0074) along with doxycycline hyclate (2 μg mL^-1^). Day 2 media was N2+SB/XAV/LDN/doxycycline hyclate and differentiation media were as previously described. On day 3, cell differentiation was continued in neurobasal media (Life Technologies, Inc.) supplemented with B27 (50X, Thermo Fisher Scientific), brain-derived neurotrophic factor (BDNF), ciliary neurotrophic factor (CNTF), glial cell-derived neurotrophic factor (GDNF) (R&D Systems 248-BD/CF, 257-NT/CF, and 212-GD/CF at 10 ng mL^-1^) and doxycycline hyclate (2 μg mL^-1^). NPC were harvested on day 4 and ExN on day 31 for all applications.

### Differentiation of ESC to InN

Human ESC were differentiated to InN as previously described [25]. Briefly, cells were cultured in a medium consisting of DMEM/F12 and Neurobasal (1:1 ratio) supplemented with N2 (1:100) and B27 without vitamin A (1:50), with the Rock inhibitor Y-27632 (10 μM, Tocris Biochem) from day 0 to day 2. On day 4, cell were re-plated in DMEM/F12 and Neurobasal (1:1 ratio) with N2 supplement (1:200) and B27 without vitamin A (1:100) on plasticware coated with polyornithine (PO), fibronectin (FN), and laminin (lam). From day 0 to day 9, the medium included SB431542 (10 μM, Tocris) and Noggin (200 ng/ml, R&D) for neuralization, along with the patterning factors SHH-C24II (R&D, 200 ng/ml unless stated otherwise) and CT99021 (Axon Medchem). On day 11, cell clusters were dissociated into single cells with accutase and replated onto PO/FN/lam-coated plates in droplets of 5–15,000 cells/μl in Neurobasal, supplemented with B27 without vitamin A (1:50), brain-derived neurotrophic factor (BDNF) (20 ng/ml), and ascorbic acid (200 μM). From day 14 onward, db-cAMP (0.5 mM) or DAPT (2.5 μM) was added to the medium for terminal differentiation (day 42).

### Protein extraction in NPC, ExN, and InN

Total protein extract was obtained by harvesting cells and either processing them immediately or snap-freezing them on dry ice for storage at −80°C. In both cases, cell pellets were washed with PBS and resuspended in 10x packed cell volume (PCV) IP lysis buffer (Thermo Fisher Scientific), with freshly added Halt protease and phosphatase inhibitors (Thermo Fisher Scientific). After a 20 min incubation time at 4°C, cells were collected by centrifugation (16,200 g, 20 min, 4°C) and resuspended in 3x PCV lysis buffer. The concentration of the samples was quantified using the Thermo Fisher BCA protein assay and when not used immediately, samples were stored at −80°C.

### Immunoprecipitations in NPC, ExN, and InN

For each individual experiment, 2 mg of fresh protein extract from the same cell differentiation batch was incubated at 4°C overnight in the presence of 2 μg of the relevant antibody or its matching isotype IgG control. All IgG antibodies were acquired from abcam (ab172730, ab37415, ab18443). On the next day, 100 μL of Protein A/G beads (Pierce) were added to each sample and incubated at 4°C for 4 hours. Flow-through was collected and beads were washed once with 1 mL lysis buffer (Pierce) supplemented with Halt protease and phosphatase inhibitors (Thermo Fisher Scientific), and twice with PBS. Beads were resuspended in 100 μL of PBS and 10% of the volume was employed for immunoblotting (see below), after being boiled in 6xSMASH buffer (50 mM Tris HCl pH 6.8, 10% Glycerol, 2% SDS, 0.02% bromophenol blue, 1% b-mercaptoethanol) for 10 min at 95°C. The remaining volume was stored at −80°C and subsequently used for mass spectrometry analysis.

### Protein extraction and immunoprecipitations in human cortex

Immunoprecipitations in cortex were performed using either an in-gel digestion protocol [74] or the sTRAP protocol [75]. The following IP-MS datasets listed in **Table S4** were derived from the in-gel protocol: HERC1_Cortex_CNCR, MAGI2_Cortex_CNCR_2, MAGI2_Cortex_CNCR_3, SETD1A_Cortex_CNCR_2, TRIO_Cortex_CNCR_2, ZMYM2_Cortex_CNCR_2; the other Cortex datasets were derived from the sTRAP protocol.

Human cortex from six adults (three healthy individuals, three diagnosed with Alzheimer’s Disease) was homogenized using a potter and pestle (Sartorius, Göttingen, Germany; 12 strokes, 900 rpm) in either 1% Maltose-neopentyl glycol buffer for in-gel protocol or 0.5% n-Dodecyl β-D-maltoside for the sTRAP protocol, both containing 25 mm HEPES, 150 mm NaCl (pH 7.4), and proteinase inhibitor (Roche), and incubated for 1 hour at 4°C. After centrifugation at 20.000×*g*, 2 μg antibody was added to the supernatant and incubated overnight at 4°C. The antibody was captured by 40 μl protein A/G plus agarose beads (Santa Cruz). After washing four times, the beads were processed for SDS-PAGE or sTRAP.

For the in-gel digestion protocol, the beads were mixed with an SDS sample buffer and heated to 98°C for 5 min. Proteins were separated on a 10% SDS polyacrylamide gel and stained with Coomassie Blue. Each sample lane was cut into three fractions, each fraction was further cut into small gel pieces and transferred to a 96-well plate (0.45 μm filter; MultiScreen-HV 96-well filter plate from Millipore). Gel pieces were destained by sequential incubation with 100 μl 50mM NH_3_HCO_3_/50% acetonitrile and then 75 μl 100% acetonitrile. Gel pieces were subsequently incubated with MS grade endolysC/trypsin (Promega) at 37°C overnight. 100 μl 0.1% TFA in 50% acetonitrile was added to each fraction, incubated for 40 min, collected in an Eppendorf tube and dried in the SpeedVac.

For the sTRAP protocol, the beads were mixed in 50 µL 5% SDS buffer containing 100 mM Tris, pH 8 and 2 µL tris(2-carboxyethyl) phosphine (50 mM), and incubated at 65°C for 30 min in a ThermoMixer (Eppendorf, Hamburg, Germany). After mixing with 5 µL 12% phosphoric acid and 400 µL washing buffer (90% methanol and 10% 1 M Tris, pH 8), the sample was transferred to sTRAP column and washed 4 times by centrifugation at 1400×*g* for 1–2 min. Trypsin/Lys-C (Promega, Madison, WI, USA) was added to the column and incubated at 37°C overnight. The column was centrifuged for 2 min. The eluents were SpeedVac dried.

### Immunoblotting

Samples for immunoblotting were stored in 6x SMASH buffer (50 mM Tris HCl pH 6.8, 10% glycerol, 2% SDS, 0.02% bromophenol blue, 1% b-mercaptoethanol), boiled for 10 min at 95°C, separated on a NuPAGE 4-12% Bis-Tris Protein Gel (Invitrogen), and transferred onto a PVDF membrane (Life Technologies) by wet transfer (100 V for 2 hours). Membranes were blocked by incubation for one hour at room temperature in 10 mL TBS and 0.1% Tween (TBST) with 5% w/v BioRad Blotting-grade Blocker. Blots were incubated overnight at 4°C with the primary antibody, washed 3 times for 10 min with TBST and incubated for 45 min with secondary antibody conjugated to horseradish peroxidase. After washing 3 times for 5 min with TBST, bands were visualized using SuperSignal^TM^ West Femto Maximum Sensitivity Substrate (Thermo Fisher Scientific). All primary antibodies used in this study are listed in **Table S4** and **S7**.

### Single-nucleus RNA sequencing

Samples for snRNA-seq were processed in three batches as indicated in **Table S1**. Nuclei were isolated from frozen cell pellets following the 10x Genomics Demonstrated Protocols for Isolation of Nuclei for Single Cell RNA Sequencing (CG000124 Rev F) with minor modifications. Briefly, based on the pellet size, between 1.0 and 2.5 ml of cold lysis buffer containing 10 mM Tris-HCl (Sigma, T2788), 10 mM NaCl (Invitrogen, AM9760G), 3 mM MgCl_2_ (Sigma, M1028-100ML) and 0.005% Nonidet P40 (Sigma, 74385) was added into the frozen cell pellet containing tube, and mixed well by pipetting with a wide-bore tip until homogenized. After incubation on ice for 5 minutes, the lysate was spun down at 500 g and 4°C for 5 minutes. After the supernatant removal, 1 ml of cold wash buffer containing 1X PBS (Invitrogen, AM9625) with 1.0% BSA (Miltenyi Biotec,130-091-376) and 0.2 U/µl RNase Inhibitor (Takara, 2313A) was used to resuspend the nuclei pellet. The nuclei suspension was spun down at 500 g and 4°C for 5 minutes followed by the supernatant removal. The washing step was repeated two more times. The final pellet was resuspended in 1 ml cold wash buffer and passed through a 35 µm blue cap filter (Thermo Fisher Scientific, 08-771-23) into a FACS tube. The nuclei were then counted using the Cellometer K2 with the AO stain (Nexcelom Bioscience, CS1-0108-5ML). Any samples that exhibited clumping of the nuclei during imaging were filtered again with a 20 µm filter (Sysmex, 04-004-2325) into a FACS tube and recounted with the same system. The nuclei of each sample were loaded onto a channel of a Chromium™ NextGEM Chip G (10x Genomics, PN-1000120) aiming for recovery of 6,000 nuclei per channel. The snRNA-seq libraries were prepared with the Chromium™ NextGEM Single Cell 3’ GEM, Library & Gel Bead Kit v3.1 (10X Genomics, PN-1000121) following the manufacturer’s protocol. The libraries were pooled based on molar concentrations and sequenced on a NextSeq 500 flow cell (Illumina) or a NovaSeq SP1 flow cell (Illumina) with 28 bases for read1, 55 bases for read2 and 8 bases for Index read1.

### GSK3 inhibition experiment

iPSC were differentiated into ExN as detailed above. At day 30.5 of differentiation, media was supplemented with BRD7355 (CDoT, Broad Institute) at a concentration of 500 nM in 0.1% DMSO; or with BRD7355 inactive enantiomer at 500 nM in 0.1% DMSO. After 12 hours, cells were harvested and processed for phosphoproteomics as described below.

### Sample collection for whole-cell proteomic and phosphoproteomic profiling

Samples were obtained by harvesting cells and snap-freezing them on dry ice for storage at - 80°C. Cells were first pelletted (200 g, 20 min, RT), then washed twice with PBS (16,200 g, 5 min, RT). For proteomic profiling of differentiating ExN, biological duplicates each containing ∼20 million cells were collected at days 0, 4, 7, 21, 31, and 51 of NGN2-mediated differentiation protocol, as described above. Similarly, for phosphoproteomic profiling of the GSK3 inhibition experiment, ∼20 million cells were collected in duplicate at day 31 of the protocol.

For proteomic profiling of the 22q11.2del carrier lines vs. controls, ∼10 million cells were collected in duplicate at days 0, 4, and 31 of the differentiation protocol. As these samples had to be analyzed in multiple MS batches, we also generated an anchor sample by pooling all samples in equal amounts based on estimated cell counts. The anchor sample was then included in all batches to facilitate downstream data normalization.

### Proteomic profiling of differentiating ExN and 22q11.2del vs. control experiments

#### Sample preparation for mass spectrometry

Mass spectrometry analysis of collected cell pellets were performed at the Thermo Fisher Center for Multiplexed Proteomics. Samples for protein analysis were prepared essentially as previously described [76]. Proteomes were extracted using a buffer containing 200 mM EPPS pH 8.5, 8M urea, and protease inhibitors. Following lysis, each sample was reduced with 5 mM TCEP. Cysteine residues were alkylated using 10 mM iodoacetamide for 20 minutes at RT in the dark. Excess iodoacetamide was quenched with 10 mM DTT. 100 µg of each proteome was precipitated and re-solubilized in 200 mM EPPS pH 8.5. Samples were digested with Lys-C (1:100) overnight at RT and subsequently with trypsin (1:100) for 6 hours at 37°C. Anhydrous acetonitrile was added to each sample to achieve a final concentration of 33% acetonitrile. 50 µg of peptides from each sample were labeled with ∼150 µg of TMTPro reagents (Thermo Fisher Scientific) for 2 hours at room temperature. Labeling reactions were quenched with 0.5% hydroxylamine and acidified with formic acid. Acidified peptides were combined and desalted by Sep-Pak (Waters). Peptides were eluted in 70% acetonitrile, 1% formic acid and dried by vacuum centrifugation.

#### Basic pH reversed-phase separation (BPRP)

TMT labeled peptides corresponding to the total proteome were solubilized in 5% ACN/10 mM ammonium bicarbonate, pH 8.0 and 300 µg of TMT labeled peptides was separated by an Agilent 300 Extend C18 column (3.5 mm particles, 4.6 mm ID and 250 mm in length). An Agilent 1260 binary pump coupled with a photodiode array (PDA) detector (Thermo Fisher Scientific) was used to separate the peptides. A 45 minute linear gradient from 10% to 40% acetonitrile in 10 mM ammonium bicarbonate pH 8.0 (flow rate of 0.6 mL/min) separated the peptide mixtures into a total of 96 fractions (36 seconds). A total of 96 Fractions were consolidated into 24 samples in a checkerboard fashion, acidified with 20 µL of 10% formic acid and vacuum dried to completion. 24 fractions were desalted via Stage Tips and re-dissolved in 5% FA/ 5% ACN for LC-MS3 analysis.

#### Liquid chromatography separation and tandem mass spectrometry (LC-MS3) for differentiating ExN experiment

Total proteome data were collected on an Orbitrap Eclipse mass spectrometer (Thermo Fisher Scientific) coupled to a Proxeon EASY-nLC 1200 LC pump (Thermo Fisher Scientific). Fractionated peptides were separated using a 180 min gradient at 500 nL/min on a 35 cm column (i.d. 100 μm, Accucore, 2.6 μm, 150 Å) packed in-house. MS1 data were collected in the Orbitrap (120,000 resolution; maximum injection time 50 ms; AGC 1 × 10^5^). Charge states between 2 and 5 were required for MS2 analysis, and a 90 s dynamic exclusion window was used. MS2 scans were performed in the ion trap with CID fragmentation (isolation window 0.5 Da; Ion Trap Scan Rate - Rapid; NCE 35%; maximum injection time 35 ms; AGC 1.5 × 10^4^). An on-line real-time search algorithm (Orbiter) was used to trigger MS3 scans for quantification [77]. MS3 scans were collected in the Orbitrap using a resolution of 50,000, NCE of 45%, maximum injection time of 200 ms, and AGC of 1.5 × 10^5^. The close out was set at two peptides per protein per fraction [77].

#### Liquid chromatography separation and tandem mass spectrometry (LC-MS3) for 22q11.2del vs. control experiment

Total proteome data were collected on an Orbitrap Fusion Lumos mass spectrometer (Thermo Fisher Scientific) coupled to a Proxeon EASY-nLC 1000 LC pump (Thermo Fisher Scientific). Fractionated peptides were separated using a 120 min gradient at 600 nL/min on a 35 cm column (i.d. 100 μm, Accucore, 2.6 μm, 150 Å) packed in-house. MS1 data were collected in the Orbitrap (120,000 resolution; maximum injection time 60 ms; AGC 10 × 10^5^). Charge states between 2 and 5 were required for MS2 analysis, and a 120 s dynamic exclusion window was used. MS2 scans were performed in the ion trap with CID fragmentation (isolation window 0.5 Da; Ion Trap Scan Rate - Rapid; NCE 35%; maximum injection time 50 ms; AGC 3 × 10^4^). An on-line real-time search algorithm (Orbiter) was used to trigger MS3 scans for quantification [77]. MS3 scans were collected in the Orbitrap using a resolution of 50,000, NCE of 45%, maximum injection time of 200 ms, and AGC of 1.5 × 10^5^. The close out was set at two peptides per protein per fraction [77].

#### Data analysis

Raw files were converted to mzXML, and monoisotopic peaks were re-assigned using Monocle [78]. Searches were performed using the Comet search algorithm against a human database downloaded from Uniprot in May 2021. We used a 50 ppm precursor ion tolerance, 1.0005 fragment ion tolerance, and 0.4 fragment bin offset for MS2 scans collected in the ion trap. TMTpro on lysine residues and peptide N-termini (+304.2071 Da) and carbamidomethylation of cysteine residues (+57.0215 Da) were set as static modifications, while oxidation of methionine residues (+15.9949 Da) was set as a variable modification.

Each run was filtered separately to 1% False Discovery Rate (FDR) on the peptide-spectrum match (PSM) level. Then proteins were filtered to the target 1% FDR level across the entire combined data set. For reporter ion quantification, a 0.003 Da window around the theoretical m/z of each reporter ion was scanned, and the most intense m/z was used. Reporter ion intensities were adjusted to correct for isotopic impurities of the different TMTpro reagents according to manufacturer specifications. Peptides were filtered to include only those with a summed signal-to-noise (SN) ≥ 100 across all TMT channels for the differentiating ExN experiment or 130 for the 22q11.2del experiment. For each protein or phosphorylation site, the filtered peptide TMTpro SN values were summed to generate protein quantification values. To control for different total protein loading within a TMTpro experiment, the summed protein quantities of each channel were adjusted to be equal within the experiment.

### Phosphoproteomic profiling of GSK3 inhibition experiment

#### Sample preparation for mass spectrometry

Mass spectrometry analysis of collected cell pellets were performed at the Thermo Fisher Center for Multiplexed Proteomics. Samples for protein analysis were prepared essentially as previously described [76]. Proteomes were extracted using a buffer containing 200 mM EPPS pH 8.5, 8M urea, 0.1% SDS and protease/phosphatase inhibitors. Following lysis, 350 µg of each proteome was reduced with 5 mM TCEP. Cysteine residues were alkylated using 10 mM iodoacetamide for 20 minutes at RT in the dark. Excess iodoacetamide was quenched with 10 mM DTT. A buffer exchange was carried out using a modified SP3 protocol [79]. Briefly, ∼3500 µg of Cytiva SpeedBead Magnetic Carboxylate Modified Particles (65152105050250 and 4515210505250), mixed at a 1:1 ratio, were added to each sample. 100% ethanol was added to each sample to achieve a final ethanol concentration of at least 50%. Samples were incubated with gentle shaking for 15 minutes. Samples were washed three times with 80% ethanol. Protein was eluted from SP3 beads using 200 mM EPPS pH 8.5 containing Lys-C (Wako, 129-02541). Samples were digested overnight at room temperature with vigorous shaking. The next morning trypsin (Thermo Fisher Scientific) was added to each sample and further incubated for 6 hours at 37° C. Acetonitrile was added to each sample to achieve a final concentration of ∼33%. Each sample was labelled, in the presence of SP3 beads, with ∼550 µg of TMTPro reagents (Thermo Fisher Scientific). Following confirmation of satisfactory labelling (>97%), excess TMT was quenched by addition of hydroxylamine to a final concentration of 0.3%. The full volume from each sample was pooled and acetonitrile was removed by vacuum centrifugation for 1 hour. The pooled sample was acidified and peptides were de-salted using a Sep-Pak 50mg tC18 cartridge (Waters). Peptides were eluted in 70% acetonitrile, 1% formic acid and dried by vacuum centrifugation.

#### Phosphopeptide enrichment and fractionation

A phosphopeptide enrichment was performed using a High-Select Fe-NTA Phosphopeptide Enrichment Kit (Thermo Fisher Scientific). Three columns were used to enrich phosphopeptides from the pooled sample. Following elution, the phosphopeptides were pooled, dried by vacuum centrifugation, and fractionated using a Pierce High pH Reversed-Phase Peptide Fractionation kit (Thermo Fisher Scientific). A total of four fractions were collected for LC-MS/MS analysis. The flow-through from the phosphopeptide enrichment was used for total proteome profiling.

#### Basic pH reversed-phase separation (BPRP) for proteome profiling

TMT labeled peptides (phosphopeptide enrichment flow-through) were solubilized in 5% acetonitrile/10 mM ammonium bicarbonate, pH 8.0 and ∼300 µg of TMT labeled peptides were separated by an Agilent 300 Extend C18 column (3.5 mm particles, 4.6 mm ID and 250 mm in length). An Agilent 1260 binary pump coupled with a photodiode array (PDA) detector (Thermo Fisher Scientific) was used to separate the peptides. A 45 minute linear gradient from 10% to 40% acetonitrile in 10 mM ammonium bicarbonate pH 8.0 (flow rate of 0.6 mL/min) separated the peptide mixtures into a total of 96 fractions (36 seconds). A total of 96 Fractions were consolidated into 24 samples in a checkerboard fashion and vacuum dried to completion. Each sample was desalted via Stage Tips and re-dissolved in 5% formic acid/ 5% acetonitrile for LC-MS3 analysis.

#### Liquid chromatography separation and tandem mass spectrometry (LC-MS3) for proteome profiling

Proteome data were collected on an Orbitrap Eclipse mass spectrometer (Thermo Fisher Scientific) coupled to a Proxeon EASY-nLC 1000 LC pump (Thermo Fisher Scientific). Fractionated peptides were separated using a 120 min gradient at 550 nL/min on a 35 cm column (i.d. 100 μm, Accucore, 2.6 μm, 150 Å) packed in-house. A FAIMS device enabled during data acquisition with compensation voltages set as −40, −60, and −80 [80]. MS1 data were collected in the Orbitrap (60,000 resolution; maximum injection time 50 ms; AGC 4 × 10^5^). Charge states between 2 and 5 were required for MS2 analysis in the ion trap, and a 120 second dynamic exclusion window was used. Top 10 MS2 scans were performed in the ion trap with CID fragmentation (isolation window 0.5 Da; Turbo; NCE 35%; maximum injection time 35 ms; AGC 1 × 10^4^). Real-time search was used to trigger MS3 scans for quantification [77]. MS3 scans were collected in the Orbitrap using a resolution of 50,000, NCE of 55%, maximum injection time of 250 ms, and AGC of 1.25 × 10^5^. The close out was set at two peptides per protein per fraction [77].

#### Liquid chromatography separation and tandem mass spectrometry (LC-MS2) for phosphoproteome profiling

Phosphorylation data were collected on an Orbitrap Eclipse mass spectrometer (Thermo Fisher Scientific) coupled to a Proxeon EASY-nLC 1000 LC pump (Thermo Fisher Scientific). Fractionated peptides were separated using a 120 min gradient at 525 nL/min on a 35 cm column (i.d. 100 μm, Accucore, 2.6 μm, 150 Å) packed in-house. A FAIMS device enabled during data acquisition with compensation voltages set as −40, −60, and −80 V for the first shot and −45 and −65 V for the second shot [80]. MS1 data were collected in the Orbitrap (120,000 resolution; maximum injection time set to auto; AGC 4 × 10^5^). Charge states between 2 and 5 were required for MS2 analysis, and a 120 second dynamic exclusion window was used. Cycle time was set at 1 second. MS2 scans were performed in the Orbitrap with HCD fragmentation (isolation window 0.5 Da; 50,000 resolution; NCE 36%; maximum injection time 300 ms; AGC 2 × 10^5^).

#### Data analysis

Raw files were converted to mzXML, and monoisotopic peaks were re-assigned using Monocle [78]. Searches were performed using the Comet search algorithm against a human database downloaded from Uniprot in May 2021. We used a 50 ppm precursor ion tolerance, 1.0005 fragment ion tolerance, and 0.4 fragment bin offset for MS2 scans collected in the ion trap, and 0.02 fragment ion tolerance; 0.00 fragment bin offset for MS2 scans collected in the Orbitrap. TMTpro on lysine residues and peptide N-termini (+304.2071 Da) and carbamidomethylation of cysteine residues (+57.0215 Da) were set as static modifications, while oxidation of methionine residues (+15.9949 Da) was set as a variable modification. For phosphorylated peptide analysis, +79.9663 Da was set as a variable modification on serine, threonine, and tyrosine residues. Each run was filtered separately to 1% False Discovery Rate (FDR) on the peptide-spectrum match (PSM) level. Then proteins were filtered to the target 1% FDR level across the entire combined data set. Phosphorylation site localization was determined using the AScorePro algorithm [81]. For reporter ion quantification, a 0.003 Da window around the theoretical m/z of each reporter ion was scanned, and the most intense m/z was used. Reporter ion intensities were adjusted to correct for isotopic impurities of the different TMTpro reagents according to manufacturer specifications. Peptides were filtered to include only those with a summed signal-to-noise (SN) ≥ 120 across all TMT channels. An extra filter of an isolation specificity (“isolation purity”) of at least 0.5 in the MS1 isolation window was applied for the phosphorylated peptide analysis. For each protein or phosphorylation site, the filtered peptide TMTpro SN values were summed to generate protein or phosphorylation site quantification values. The signal-to-noise (S/N) measurements of peptides assigned to each protein were summed (for a given protein). These values were normalized so that the sum of the signal for all proteins in each channel was equivalent thereby accounting for equal protein loading. The resulting normalization factors were used to normalize the phosphorylation sites, again accounting for equal protein loading.

### Mass spectrometry for IP-MS experiments

Mass spectrometry analysis of IP samples in NPC, ExN, and InN was performed at the Harvard Center for Mass Spectrometry (Harvard) or Taplin Mass Spectrometry Facility (Taplin); analysis for IPs in human cortex was performed at Center for Neurogenomics and Cognitive Research (CNCR; see **Table S4** for details).

#### Harvard

IP samples were stored in PBS buffer on beads. PBS was removed and samples were dissolved in 50 μL of 50 mM TEAB, followed by trypsin (Promega) digestion for 3 hr at 38°C. Digested samples were dried to 20 µL and 10 μL of each sample was injected in the mass spectrometer. LC-MS/MS was performed on a Lumos Tribrid Orbitrap Mass Spectrometer (Thermo Fisher Scientific) equipped with Ultimate 3000 (Thermo Fisher Scientific) nano-high-performance liquid chromatography. Peptides were separated onto a 150-μm inner diameter microcapillary trapping column, packed with ∼2 cm of C18 Reprosil resin (5 μm, 100 A□, Dr. Maisch GmbH, Germany), followed by separation on a 50-cm analytical column (PharmaFluidics, Ghent, Belgium). Separation was achieved by applying a gradient from 5 to 27% acetonitrile in 0.1% formic acid for > 90 min at 200 nL min^-1^. Electrospray ionization was enabled by applying a voltage of 2 kV using a home-made electrode junction at the end of the microcapillary column and sprayed from metal tips (PepSep, Denmark). MS survey scan was performed in the Orbitrap, in a range of 400–1800 m/z at a resolution of 60,000, followed by the selection of the 20 most intense ions (TOP20) for CID-MS2 fragmentation in the ion trap using a precursor isolation width window of 2 m/z, automatic gain control setting of 10,000, and a maximum ion accumulation of 100 ms. Singly charged ion species were not subjected to collision-induced dissociation fragmentation. Normalized collision energy was set to 35 V and an activation time of 10 ms. Ions within a 10 ppm m/z window around ions selected for MS2 were excluded from further selection for fragmentation for 60 s.

Mass spectra were analyzed using Proteome Discoverer (v2.4; Thermo Fisher Scientific). Assignment of MS/MS spectra was performed using the Sequest HT algorithm by searching the data against the UniProt human reference proteome, including isoforms (Swiss-Prot, release 2019_01) as well as other known contaminants such as human keratins and common laboratory contaminants. Sequest HT searches were performed using a 10 ppm precursor ion tolerance and requiring each peptide’s N/C termini to adhere with trypsin protease specificity, while allowing up to two missed cleavages. CID-MS2 spectra were searched with 0.5 Da ion tolerance for fragmentation. Methionine oxidation (+15.99492 Da) was set as variable modification. An MS2 spectra assignment FDR of 1% was applied to both proteins and peptides using the Percolator target-decoy database search. Label-free quantitation was performed to generate protein-level quantification reports.

#### Taplin

IP samples on beads were washed at least five times with 100 µl 50 mM ammonium bicarbonate then 5 µl (200ng/ul) of modified sequencing-grade trypsin (Promega, Madison, WI) was spiked in and the samples were placed in a 37°C room overnight. The samples were then centrifuged or placed on a magnetic plate if magnetic beads were used and the liquid removed. The extracts were then dried in a speed-vac (∼1 hr). Samples were then re-suspended in 50 µl of HPLC solvent A (2.5% acetonitrile, 0.1% formic acid) and desalted by STAGE tip [82].

On the day of analysis the samples were reconstituted in 10 µl of HPLC solvent A. A nano-scale reverse-phase HPLC capillary column was created by packing 2.6 µm C18 spherical silica beads into a fused silica capillary (100 µm inner diameter x ∼30 cm length) with a flame-drawn tip [83]. After equilibrating the column 4 µl of each sample was loaded via a Famos auto sampler (LC Packings, San Francisco CA) onto the column. A gradient was formed and peptides were eluted with increasing concentrations of solvent B (97.5% acetonitrile, 0.1% formic acid). As peptides eluted they were subjected to electrospray ionization and then entered into an LTQ Orbitrap Velos Pro, Exploris 480, or Fusion Lumos mass spectrometer (Thermo Fisher Scientific, Waltham, MA). Peptides were detected, isolated, and fragmented to produce a tandem mass spectrum of specific fragment ions for each peptide.

Peptide sequences (and hence protein identity) were determined by matching the UniProt human protein database (release 2023_01) with the acquired fragmentation pattern using Sequest (Thermo Fisher Scientific, Waltham, MA) [84]. The database included a reversed version of all the sequences and the data were filtered to between 1-2% peptide false discovery rate. Protein quantification was performed using GFY Core Version 3.8 (Harvard University, Cambridge, MA).

#### CNCR

The tryptic peptides from in-gel digestion and sTRAP protocols were transferred to the Evotip Pure (Evosep, Odense C, Denmark), and separated on the EV1137 reverse-phase column (Evosep, Odense C, Denmark) using the Evosep One LC system (Evosep, Odense C, Denmark) with the 30 samples per day program. Peptides were analyzed by the TimsTOF Pro 2 mass spectrometer (Bruker Daltonics, Bremen, Germany). The MS was operated with scan range 100–1700 m/z, ion mobility 0.6 to 1.6LVs/cm^2^, ramp time 100 ms, accumulation time 100 ms, and collision energy decreasing linearly with the inverse precursor ion mobility from 59 eV at 1.6 Vs/cm^2^ to 20 eV at 0.6 Vs/cm^2^. For the operation in dia-PASEF mode [75], each cycle took 1.8 s and consisted of 1 MS1 full scan and 16 dia-PASEF scans. Each dia-PASEF scan contained two dia-PASEF isolation windows, in total covering 400–1201 m/z (1 Th window overlap) and ion mobility 0.6 to 1.43LVs/cm^2^. The dia-PASEF raw data were processed with dia-NN (v1.8.1). An in-silico spectral library was generated from the Uniprot human proteome (UP000005640_9606, release 2023-03) and was used for database search with the following settings: trypsin/P digestion and at most 1 missed cleavage, methionine oxidation was enabled, maximum number of variable modifications was set to 1, precursor charge range was set to 2-4, precursor length range 6-30, precursor m/z range 300-1400, fragment ion m/z 200-1800, both MS1 and MS2 mass accuracy were set to 10 ppm. All other settings were left as default.

### snRNA-seq data analysis

The snRNA-seq data were processed with CellRanger v6.1.2 [85] using the refdata-gex-GRCh38-2020-A reference from 10x Genomics, with the flags --expect-cells=10000 -- localmem=64 --nosecondary --chemistry=SC3Pv3 --include-introns. Nuclei with <300 genes expressed were excluded from downstream analysis. During QC, three iPSC samples and one ExN sample showed elevated levels of mitochondrial gene expression (**Table S1**), which could indicate extranuclear contamination during nuclei isolation. To mitigate this issue, we regressed out the percentage of mitochondrial genes as a technical variable when performing data normalization using the SCTransform method in Seurat [86] (v5.0.3). Furthermore, we repeated downstream analysis with vs. without filtering of nuclei with ≥5% mitochondrial genes to confirm that the inclusion of these nuclei did not influence the findings described in this study. To integrate data generated across different batches (i.e., 10x runs performed on different dates; **Table S1**), we ran STACAS [87] (v2.2.2) using 3000 anchor features, 30 dimensions, and cell type labels (iPSC, NPC, ExN, InN) for semi-supervised alignment, followed by UMAP dimensional reduction on the integrated Seurat object for downstream data visualization.

We used ProjecTILs [26] (v3.3.0) to map the stem cell-derived neurons onto a human cortex reference. We first created a reference map using snRNA-seq data for “All Cells” from Velmeshev et al. [27] (https://cells.ucsc.edu/?ds=pre-postnatal-cortex). Next, we separately projected each batch of iPSC, NPC, ExN, and InN onto the reference map, then merged the results across all batches in combined projection plots for visualization.

### Whole proteome data analysis

To analyze data from the differentiating ExN proteomic profiling experiment, we first normalized protein-level quantities from 12 samples (6 differentiation time points in duplicate; analyzed as a single TMT 12-plex) so that the total signal from each TMT-channel was the same. The abundance values for each protein were then scaled to sum to 100 across all samples. We removed proteins that were not assigned a gene symbol. As proteins were analyzed based on their gene symbols in downstream analysis, for proteins where multiple Uniprot IDs were assigned to the same gene symbol, only one value was kept, based on the following criteria: (1) keep ‘sp’ (UniProtKB/Swiss-Prot) annotated proteins over ‘tr’ (UniProtKB/TrEMBL) annotated proteins, (2) for cases with multiple ‘sp’ annotated proteins or no ‘sp’ and multiple ‘tr’ annotated proteins, keep entry with highest signal to noise values. We performed principal component analysis to evaluate major drivers of changes in cellular proteome across neuronal differentiation. We also examined expression of cell type marker genes [23] to validate successful neuronal differentiation.

We performed k-means clustering of scaled protein expression across neuronal differentiation. Optimal number of clusters (k=5) was determined by the elbow method. Biological functions of proteins in each cluster were evaluated by gene set enrichment analysis of GO Biological Process terms [88] (as curated in Genoppi [24], development branch v1.1.0) using one-tailed hypergeometric tests and all proteins detected in this experiment as the background population.

See “*22q11.2del proteomic analysis*” and “*GSK3 inhibition phosphoproteomic analysis*” below for analysis of other whole-cell proteomic profiling datasets.

### Gene and protein expression plots

UMAP plots showing snRNA-seq expression (normalized log counts per million [CPM]) of genes of interest were generated using Seurat [86] (v5.0.3) and the human cortex reference map or the STACAS-integrated dataset from stem cell-derived cell types, described in “*snRNA-seq data analysis”* above. In addition, we calculated pseudobulk expression for the iPSC, NPC, and ExN samples (using the AggregateExpression function in Seurat) to generate an expression heat map for the SCZ index genes. We also generated an analogous protein expression heat map using scaled abundance values from the whole proteome dataset described in “*Whole proteome data analysis”* above.

### IP-MS data analysis

Starting from protein-level quantification data, we performed the following for each IP-MS dataset (consisting of bait vs. IgG control IPs in triplicates): (1) log_2_ transform and median normalize protein intensities in each sample; (2) remove contaminants and proteins with no human gene name, detected with < 2 unique peptides, or detected in < 2 bait IP samples; (3) impute missing values for the remaining proteins in each sample by drawing from a normal distribution with mean of μ - 1.8σ and standard deviation of 0.3σ, where μ and σ are the mean and standard deviation of the observed protein intensities, respectively [89]; (4) calculate replicate correlations among bait and control IPs; (5) perform limma-based [90] two-tailed two-sample moderated t-test as implemented in Genoppi [24] (development branch v1.1.0), to calculate enrichment statistics (log_2_ fold change [FC], p-value, and false discovery rate [FDR]) of each protein in bait vs. control IPs; (6) define proteins with log_2_ FC > 0 and FDR ≤ 0.1 as significant interactors of the bait protein. We excluded IP-MS datasets in which the bait protein itself was not positively enriched at FDR ≤ 0.1 from further analysis. The remaining 56 datasets are summarized in **Tables S4** and **S5**.

### Systematic comparison of IP-MS datasets

To systematically compare the IP-MS datasets generating using different bait proteins, cellular contexts, and MS facilities, we first calculated the following summary statistics for each dataset: (1) number of analyzed proteins; (2) mean Pearson correlation of log_2_ protein abundance among bait or control IP replicates; (3) proportion of proteins with imputed values among bait IPs, control IPs, or both; (4) number of significant interactors of the bait protein; (5) number of significant interactors that are in InWeb [16]; (6) overlap enrichment p-value between significant vs. InWeb interactors, calculated using a one-tailed hypergeometric test; (7) log_2_ FC and FDR of bait protein; (8) median and maximum log_2_ FC and FDR of significant interactors. We subsetted the IP-MS datasets by cellular context or MS facility, then compared the statistics of each group using violin plots and two-tailed Wilcoxon rank-sum tests.

Next, we calculated several similarity metrics for pairwise comparison of IP-MS datasets, including: (1) Jaccard index quantifying overlap of detected proteins or significant interactors; (2) log_2_ FC or signed (based on log_2_ FC) −log_10_ FDR correlations of proteins detected in both datasets; (3) one-tailed hypergeometric p-value indicating enrichment of interactor overlap; (4) Spearman correlation based on GO term enrichment p-values, which represents pathway-level overlap. To calculate the last metric, we first performed one-tailed hypergeometric tests to calculate overlap enrichment between interactors identified in each IP-MS dataset and 12,214 GO Biological Process terms (as curated in Genoppi [24], development branch v1.1.0), using all Ensembl (release 109) [91] protein-coding genes (minus the baits) as the background population. We identified 370 GO terms that were significantly enriched in ≥1 IP-MS dataset (Benjamini-Hochberg FDR < 0.05), then calculated the Spearman correlation for each pair of IP-MS datasets using the enrichment p-values for these GO terms.

### Generating PPI networks across cellular contexts

To generate the unified PPI network across cellular contexts, we aggregated unique interactions (bait-interactor pairs) and interactors identified across all IP-MS datasets in **Tables S9** and **S10**, respectively. These tables summarize the number and name of cellular contexts (NPC, ExN, InN, Cortex) that detected each interaction or interactor. To define PPI sub-networks, we parsed the interactions table based on the cellular context and/or bait protein annotations. In particular, the NPC-Cortex, ExN-Cortex, and InN-Cortex sub-networks were defined using overlap of interactions between the contexts (not overlap of interactors).

For the interactions table, we also annotated any overlap with known interactions in PPI databases/datasets curated in Genoppi (development branch v1.1.0), including InWeb [16], BioPlex [29], iRefIndex [30], and STRING [31]. For the interactors table, we added the following annotations: (1) whether the interactor had been implicated in genetic studies of SCZ (Supplementary Table 5 of Singh et al. [3], Supplementary Tables 3 and 12 of Trubetskoy et al. [2]), ASD, and DD (Supplementary Tables 11 and 15 of Fu et al. [34]); (2) *postmortem* brain cell types in which the interactor was identified as a differentially expressed gene in SCZ cases vs. controls (Supplementary Table 4 of Ruzicka et al.[7]); (3) whether the interactor was identified as a differentially expressed protein in 22q11.2del vs. control ExN (see “*22q11.2del proteomic analysis*” below); (4) whether the interactor contains differentially expressed phosphopeptides identified in GSK3 inhibitor-treated vs. control ExN (see “*GSK3 inhibition phosphoproteomic analysis*” below); (5) known drugs targeting the interactor as curated in the Open Targets Platform [46] (v23.12).

To visualize the unified PPI network across all cellular contexts, we generated a network graph using the igraph (v1.5.1) and qgraph (v1.9.8) R packages, in which vertices represent index or interactor proteins and edges represent observed PPI.

### GTEx tissue enrichment analysis

We obtained tissue specificity scores from Finucane et al. [22] (https://console.cloud.google.com/storage/browser/broad-alkesgroup-public-requester-pays/LDSCORE/LDSC_SEG_ldscores/tstats) and defined top 10% of genes in each tissue (or brain region) as the tissue-specific gene set. For each PPI network, we performed a one-tailed hypergeometric test to calculate overlap enrichment between the network genes vs. each gene set, using all Ensembl (release 109) [91] protein-coding genes as the background population. In this and all subsequent network enrichment analyses described below, the index genes for each network were always excluded from the analysis to avoid biasing the enrichment signals.

### SynGO enrichment analysis

SynGO enrichment analysis was performed using the SynGO browser [33] (https://syngoportal.org; 20231201 release). The PPI network genes were compared against the SynGo “brain expressed” background in one-tailed Fisher’s exact tests to identify overrepresented synaptic terms.

### SCHEMA social Manhattan plot

We generated a “social Manhattan plot” to visualize the intersection between the PPI network and SCZ rare variant risk genes prioritized in SCHEMA. SCZ association p-values (y-axis) of the index genes and their interactors were extracted from Supplementary Table 5 of Singh et al. [3]; only interactors with *P* < 6.94e-4 (corresponding to FDR ≤ 0.25) were included. The GRCh38 genomic coordinates (x-axis) of these genes were retrieved from Ensembl (release 112) [91]. Plotted links connecting a pair of genes indicate observed PPI in our data.

### Rare variant enrichment analysis

Gene-based association statistics were obtained from exome sequencing studies of SCZ (“P meta” column in Supplementary Table 5 of Singh et al. [3]), ASD, and DD (“FDR_TADA_ASD” and “FDR_TADA_DD” columns in Supplementary Table 11 of Fu et al. [34]). For each phenotype, we performed one-tailed Kolmogorov-Smirnov tests to assess whether the PPI network genes have more significant association statistics compared to other genes in the background. For analyzing the combined networks across cellular contexts (**Table S15**), we used all protein-coding genes included in each genetic study as the background; for analyzing the ExN sub-networks (**Table S17**), we used genes detected in the proteome of differentiating ExN as the background (see “*Whole proteome data analysis”* above).

### Common variant enrichment analysis

Variant-to-gene annotation files generated from human fetal or adult brain Hi-C data [37, 42] were downloaded from the H-MAGMA [36] GitHub (https://github.com/thewonlab/H-MAGMA). We mapped the gene IDs in the annotation files to gene names using Ensembl (release 109) [91]. GWAS summary statistics for SCZ [2], ADHD [38], ASD [39], BIP [40], and MDD [41] were obtained from the Psychiatric Genomics Consortium (https://www.med.unc.edu/pgc/download-results/). For each GWAS phenotype, we computed gene-based p-values using the SNP-wise mean model in MAGMA [35] (v1.09), the 1000 Genomes [92] (phase3) EUR panel, and the H-MAGMA annotation files. Next, we performed MAGMA gene-set analysis for each PPI network, computing a one-tailed p-value to assess whether the network genes are more significantly associated with the GWAS phenotype compared to other genes in the background. For analyzing the combined networks across cellular contexts (**Table S16**), we used all protein-coding genes in the H-MAGMA annotation files as the background; for analyzing the ExN sub-networks (**Table S18**), we further filtered for genes detected in the proteome of differentiating ExN as the background (see “*Whole proteome data analysis”* above).

### SCZ DEG enrichment analysis

Differentially expressed genes (DEGs) in SCZ cases vs. controls across *postmortem* prefrontal cortex cell types were obtained from Data S4 of Ruzicka et al. [7]. We performed one-tailed hypergeometric tests to calculate overlap enrichment between the PPI network genes vs. SCZ DEGs in each cell type, using all genes detected in the proteome of differentiating ExN as the background population.

### 22q11.2del#proteomic analysis

Starting with protein-level quantification data across 4 MS batches (i.e., 4 TMT 13-plexes), we first removed proteins that were not assigned a gene symbol or were detected in <2 out of the 4 plexes. As proteins were analyzed based on their gene symbol in downstream analysis, for proteins where multiple Uniprot IDs were assigned to the same gene symbol, only one value was kept, based on the following criteria: (1) keep ‘sp’ (UniProtKB/Swiss-Prot) annotated proteins over ‘tr’ (UniProtKB/TrEMBL) annotated proteins, (2) for cases with multiple ‘sp’ annotated proteins or no ‘sp’ and multiple ‘tr’ annotated proteins, keep entry with highest signal to noise values. Next, we normalized the data across plexes using an approach described in the Cancer Cell Line Encyclopedia proteomic profiling study [93]. Briefly, the summed protein quantities were adjusted to be equal across all samples within each plex and log_2_-transformed. Within each plex, protein quantity of the anchor sample was subtracted from each sample quantity and the anchor samples were removed. Finally, the mean abundance of each protein within each plex was centered at zero. We performed principal component analysis to confirm the samples did not cluster by batches/plexes after anchor normalization. We also examined expression of cell type marker genes [23] to validate successful neuronal differentiation.

Differential expression analysis between 22q11.2del vs. control iPSC, NPC, or ExN were performed using a linear mixed model in the limma R package (v3.54.2) with deletion status and differentiation day as fixed effect variables and plex as random effect variable. Two-tailed Wilcoxon rank-sum test was used to compare expression of proteins encoded by genes in the 22q11.2del region (chromosome 22, coordinates 18924718-21111383 in GRCh38.p13) in deletion vs. control samples. Gene set enrichment analysis was performed using geneSetTest function from limma R package (v3.54.2) to identify Biological Process GO terms [88] (as curated in Genoppi [24], development branch v1.1.0) differentially expressed in the 22q11.2del vs. control cells.

We performed one-tailed hypergeometric tests to calculate overlap enrichment between the PPI network genes vs. the nominally significant (*P* < 0.05) DEPs identified in iPSC, NPC, or ExN, using all genes detected in the 22q11.2del proteomic dataset as the background population.

### GSK3 inhibition phosphoproteomic analysis

We analyzed the protein-level and phosphopeptide-level quantification data from the GSK3 inhibition experiment separately. We performed log_2_ transformation on the summed intensity values for each protein or phosphopeptide, then used limma [90] (v3.54.2) to identify differentially expressed proteins or phosphopeptides (DE phosphopeptides) in GSK3 inhibitor-treated vs. control ExN at FDR < 0.05.

We performed one-tailed hypergeometric tests to calculate overlap enrichment between the PPI networks, known GSK3B substrates [50], GSK3 interactors derived from IP-MS experiment in ExN (**Table S23**), and DE phosphopeptides in ExN upon GSK3 inhibition. For all tests without the DE phosphopeptides, overlap was assessed on the gene level, using all genes detected in the proteome of differentiating ExN as the background population. For all tests including the DE phosphopeptides, overlap was assessed on the phosphopeptide level, using all phosphopeptides detected in the GSK3 inhibition experiment as the background population.

### SCZ modules analysis

We summarized results across the SCZ rare variant, SCZ common variant, SCZ DEG, 22q11.2del DEP, and GSK3 inhibition DE phosphopeptide analyses described above to assess convergence among the ExN PPI networks and define the SCZ modules. We tabulated the enrichment p-value for each network in each analysis (or minimum p-value, if multiple tests were performed across cell types and DE directions), then calculated a “summed enrichment score” that indicates the number of analyses (out of five) in which each network was at least nominally significant. We also calculated overlap enrichment p-values between pairs of networks using one-tailed hypergeometric tests and the proteome of differentiating ExN as the background. We combined these data in a network graph using igraph (v1.5.1), in which the vertex size and color scale with the size and summed enrichment score of each network, and the edge width and color scale with the number of overlapping genes and overlap significance between each pair of networks. Based on the convergence patterns in this graph, we defined three SCZ modules consisting of networks with summed enrichment scores ≥3: Module 1 consists of the GRIA3 network, Module 2 consists of the HCN4 network, and Module 3 consists of the SETD1A, TRIO, RB1CC1, AKAP11, and SRRM2 networks.

For the seven networks in the modules, we performed one-tailed hypergeometric tests to calculate overlap enrichment between the network genes and REACTOME pathways [52] (as curated in Genoppi [24], development branch v1.1.0), using all genes detected in the proteome of differentiating ExN as the background population. We used ComplexHeatmap (v2.14.0) to generate a clustered heat map showing −log_10_ enrichment p-values for the seven networks across ten REACTOME pathways, which encompass the top three enriched pathways for each network. Hierarchical clustering was performed using the default settings (“euclidean” distance and “complete” method) and the full range of −log_10_ p-values, but the maximum −log_10_ p-value was capped at 30 for color visualization in the heat map.

## Data availability

All snRNA-seq and mass spectrometry data generated in this study will be made public upon acceptance of the manuscript. Raw snRNA-seq data will be deposited at GEO; the processed data (cell-count matrix and metadata) will be available via the Single Cell Portal (https://singlecell.broadinstitute.org/single_cell). Mass spectra for the IP-MS, proteomic, and phosphoproteomic profiling experiments will be deposited at MassIVE (https://massive.ucsd.edu). Western blot images will be available on Mendeley.

## Code availability

Original code will be deposited at GitHub and Zenodo.

## Notes

### Author Declarations

Postmortem brain tissues were collected from donors with written informed consent for brain autopsy and the use of brain tissue and clinical information for research purposes. The brain donor program of the Netherlands Brain Bank (NBB) was approved by the medical ethics committee of the Vrije Universiteit (VU University) Medical Center, Amsterdam, Netherlands.

