## Supplementary_Figures for "A foundational neuronal protein network model unifying multimodal genetic, transcriptional, and proteomic perturbations in schizophrenia"

This file includes **Figures S1** to **S9**:

Figure S1. Stem cell-derived neurons model human cortical development *in vitro*.

Figure S2. Expression of SCZ index genes.

Figure S3. QC metrics of 56 IP-MS datasets across four cellular contexts.

Figure S4. Replication of identified interactions via immunoblotting and pairwise comparison of the 56 IP-MS datasets.

Figure S5. Comparison of PPI networks derived from each cellular context.

Figure S6. Genetic and transcriptional signals in the PPI network.

Figure S7. Proteomic analysis of 22q11.2del vs. control cells across neuronal differentiation.

Figure S8. Phosphoproteomic analysis of GSK3 inhibitor-treated vs. control ExN.

Figure S9. Distinct SCZ modules identified through multimodal PPI network enrichment analyses.

Separate Excel files are provided for **Tables S1** to **S27**:

Table S1. Summary of snRNA-seq profiling of stem cell-based neuronal models.

Table S2. Proteomic profiling data for differentiating ExN.

Table S3. Overlap enrichment statistics between GO Biological Process terms and protein clusters in differentiating ExN.

Table S4. Summary of 56 IP-MS datasets for 13 SCZ index proteins.

Table S5. Genoppi analysis results of 56 IP-MS datasets.

Table S6. Summary of western blot validation experiments for interactions identified via IP-MS.

Table S7. Pairwise comparison between IP-MS datasets.

Table S8. Overlap enrichment p-values between GO Biological Process terms and interactors identified in each IP-MS dataset.

Table S9. Annotation table for unique interactions in the unified PPI network.

Table S10. Annotation table for unique interactors in the unified PPI network.

Table S11. Overlap enrichment statistics between combined PPI networks derived from each cellular context.

Table S12. Overlap enrichment statistics between GTEx tissue-specific genes vs. combined PPI network from each cellular context.

Table S13. Overlap enrichment statistics between GTEx brain tissue-specific genes vs. combined PPI network from each cellular context.

Table S14. SynGO analysis results for PPI network derived from each cellular context.

Table S15. Rare variant enrichment results for combined PPI networks.

Table S16. Common variant enrichment results for combined PPI networks.

Table S17. Rare variant enrichment results for ExN PPI sub-networks.

Table S18. Common variant enrichment results for ExN PPI sub-networks.

Table S19. Overlap enrichment statistics between cell type-specific SCZ DEGs from postmortem cortex vs. ExN PPI sub-networks.

Table S20. Differential expression statistics for the 22q11.2del proteomic profiling dataset.

Table S21. Gene set enrichment statistics for DEPs in 22q11.2del iPSC, NPC, and ExN.

Table S22. Overlap enrichment statistics between DEPs in 22q11.2del iPSC, NPC, or ExN vs. ExN PPI sub-networks.

Table S23. Genoppi analysis results of GSK3B IP-MS experiment in ExN.

Table S24. Differential expression statistics for phosphopeptides detected in GSK3 inhibition experiment in ExN.

Table S25. Overlap enrichment statistics between DE phosphopeptides in GSK3-inhibited ExN vs. ExN PPI sub-networks.

Table S26. Overlap enrichment statistics between each pair of ExN index protein-specific sub-networks.

Table S27. Overlap enrichment statistics between ExN sub-networks in SCZ modules vs. REACTOME pathways.

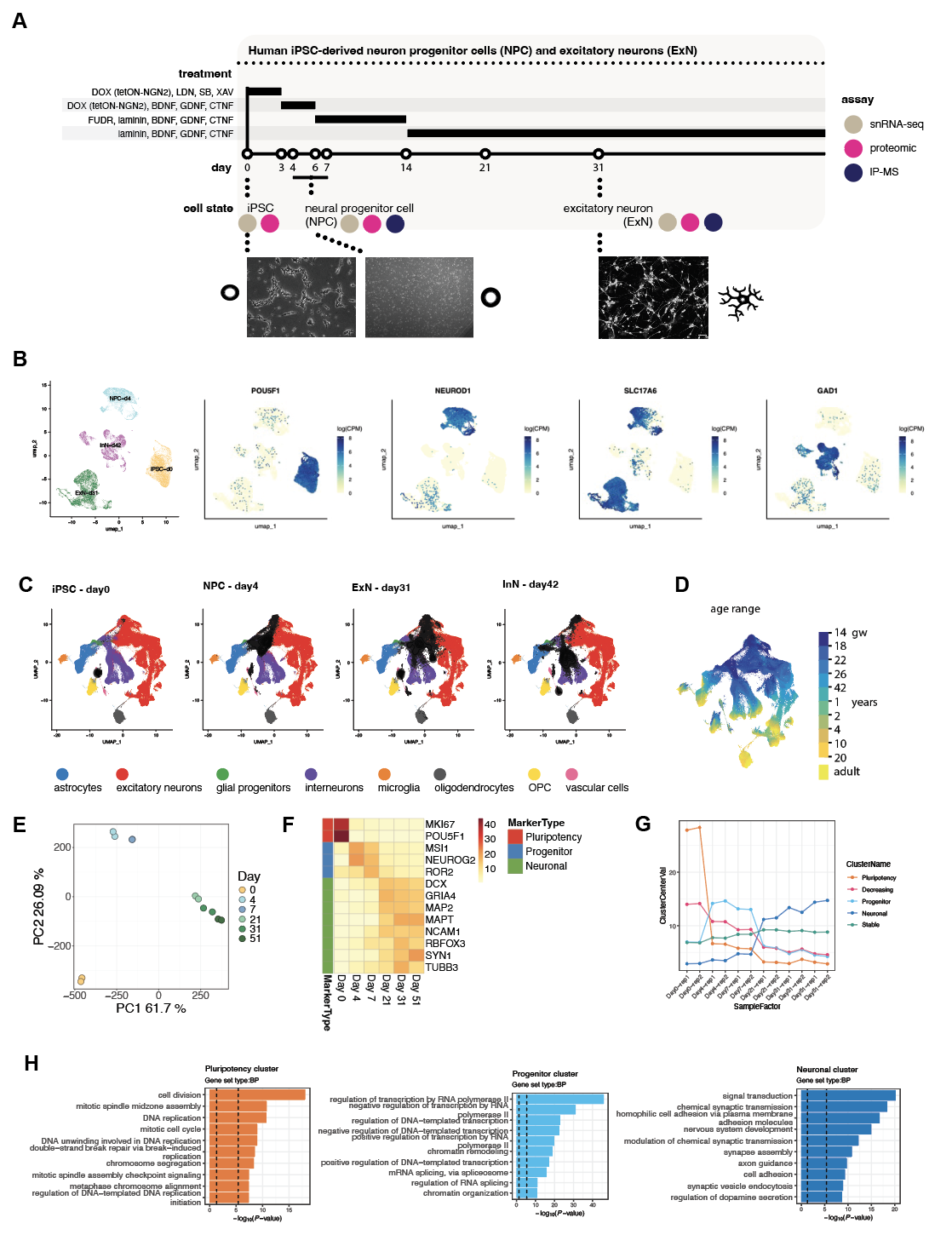

**Figure S1. Stem cell-derived neurons model human cortical development *in vitro*. (A)** Protocol to differentiate iPSC into NPC and ExN. **(B)** snRNA-seq profiling of stem cell-derived cell types (iPSC, NPC, ExN, InN) shows expected cell type marker expression: *POU5F1* for pluripotency, *NEUROD1* for neural progenitors, *SLC17A6* for excitatory neurons, and *GAD1* for inhibitory neurons. **(C)** Projections of iPSC, NPC, ExN, and InN onto a reference map of prenatal and postnatal human cortex (Velmeshev *et al*, Science, 2023) using snRNA-seq data and ProjecTILs. Colored data points in the UMAP space indicate cell lineages observed in the human cortex (see bottom legends); overlaid black triangles indicate the projected iPSC, NPC, ExN, or InN populations. **(D)** Developmental age range in the human cortex reference map from (C). gw, gestational week. **(E)** Principal component analysis of proteomic profiles of differentiating ExN. **(F)** Protein expression of cell type markers in differentiating ExN. **(G)** Average expression of protein clusters in differentiating ExN. **(H)** Top enriched gene sets (GO BP terms) for the “Pluripotency”, “Progenitor” and “Neuronal” clusters from (G). Vertical dashed lines indicate *P* < 0.05 (left) and *P* < 0.05/number of gene sets (right).

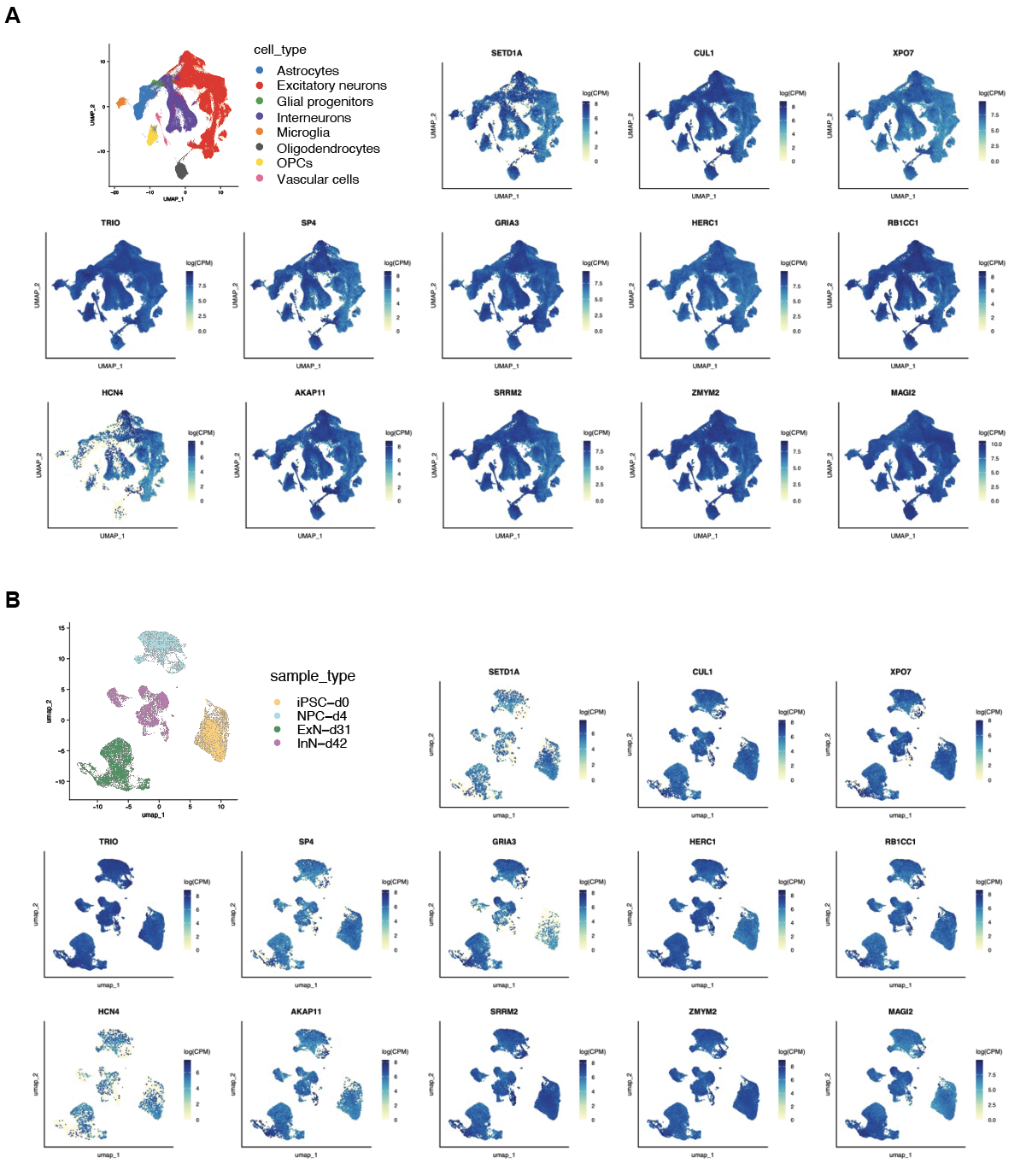

**Figure S2. Expression of SCZ index genes.** snRNA-seq expression of 13 SCZ index genes in human cortex **(A)** and stem cell-derived neurons **(B)**. CPM, counts per million.

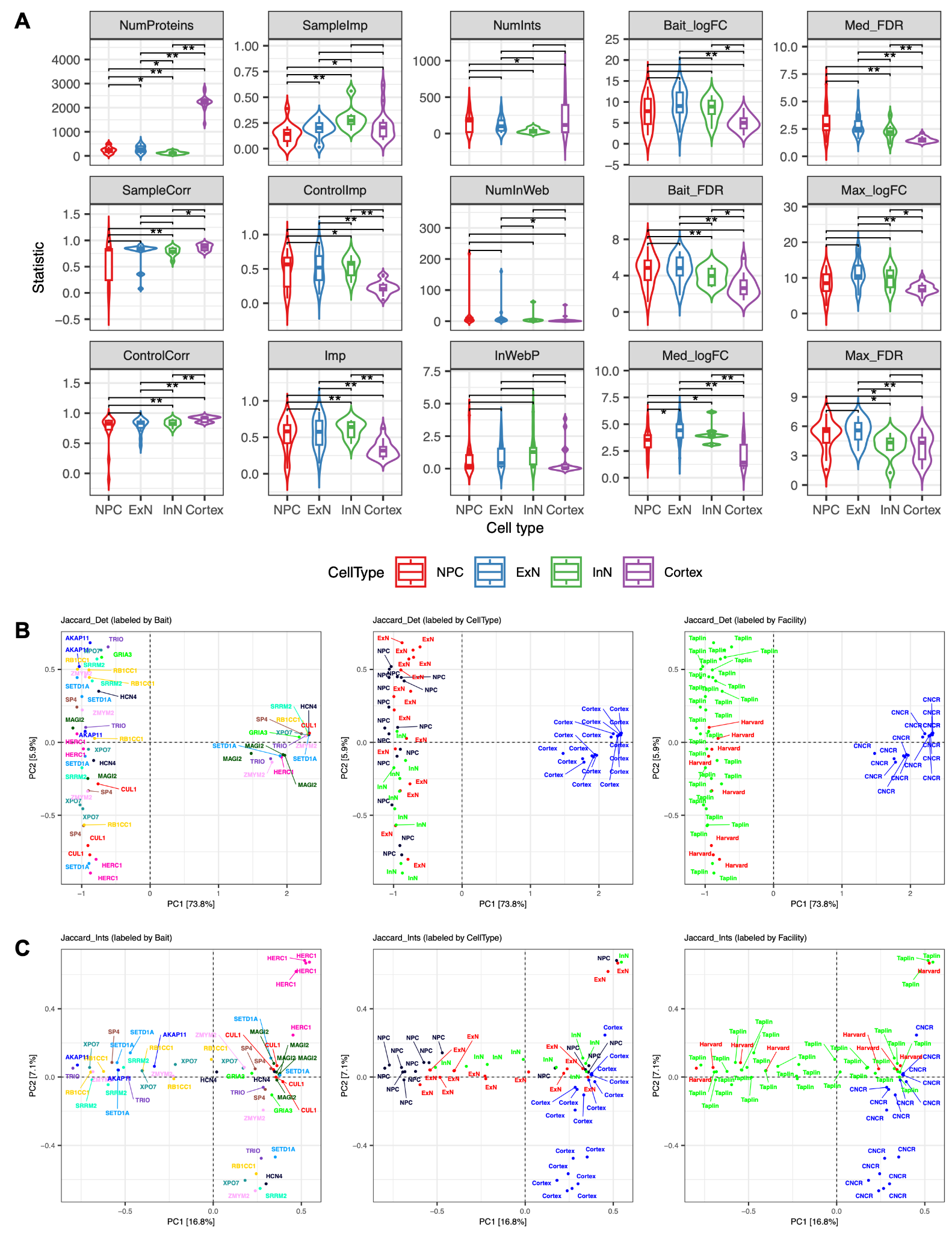

**Figure S3. QC metrics of 56 IP-MS datasets across four cellular contexts. (A)** Comparison of QC metrics across cellular contexts. NumProteins, number of analyzed proteins; SampleCorr or ControlCorr, mean log_2_ protein abundance correlation among bait or control IP replicates; SampleImp, ControlImp, or Imp, proportion of proteins with imputed values among bait IPs, control IPs, or both; NumInts, number of significant interactors of the bait protein; NumInWeb, number of significant interactors that are in InWeb; InWebP, overlap enrichment p-value between significant vs. InWeb interactors, calculated using a one-tailed hypergeometric test; Bait_logFC or Bait_FDR, log_2_ fold change (FC) or -log_10_ false discovery rate (FDR) of bait protein; Med_logFC, Med_FDR, Max_logFC, or Max_FDR, median or maximum log_2_ FC or -log_10_ FDR of significant interactors, respectively. Single asterisk indicates *P* < 0.05 and double asterisks indicate *P* < 0.005, calculated using two-tailed Wilcoxon rank-sum tests. **(B)-(C)** Principal component analysis of Jaccard index matrix quantifying overlap of detected proteins (B) or significant interactors (C) between each pair of IP-MS datasets. Each data point corresponds to an IP-MS dataset, color-coded by bait protein (left), cellular context (middle), or MS facility (right).

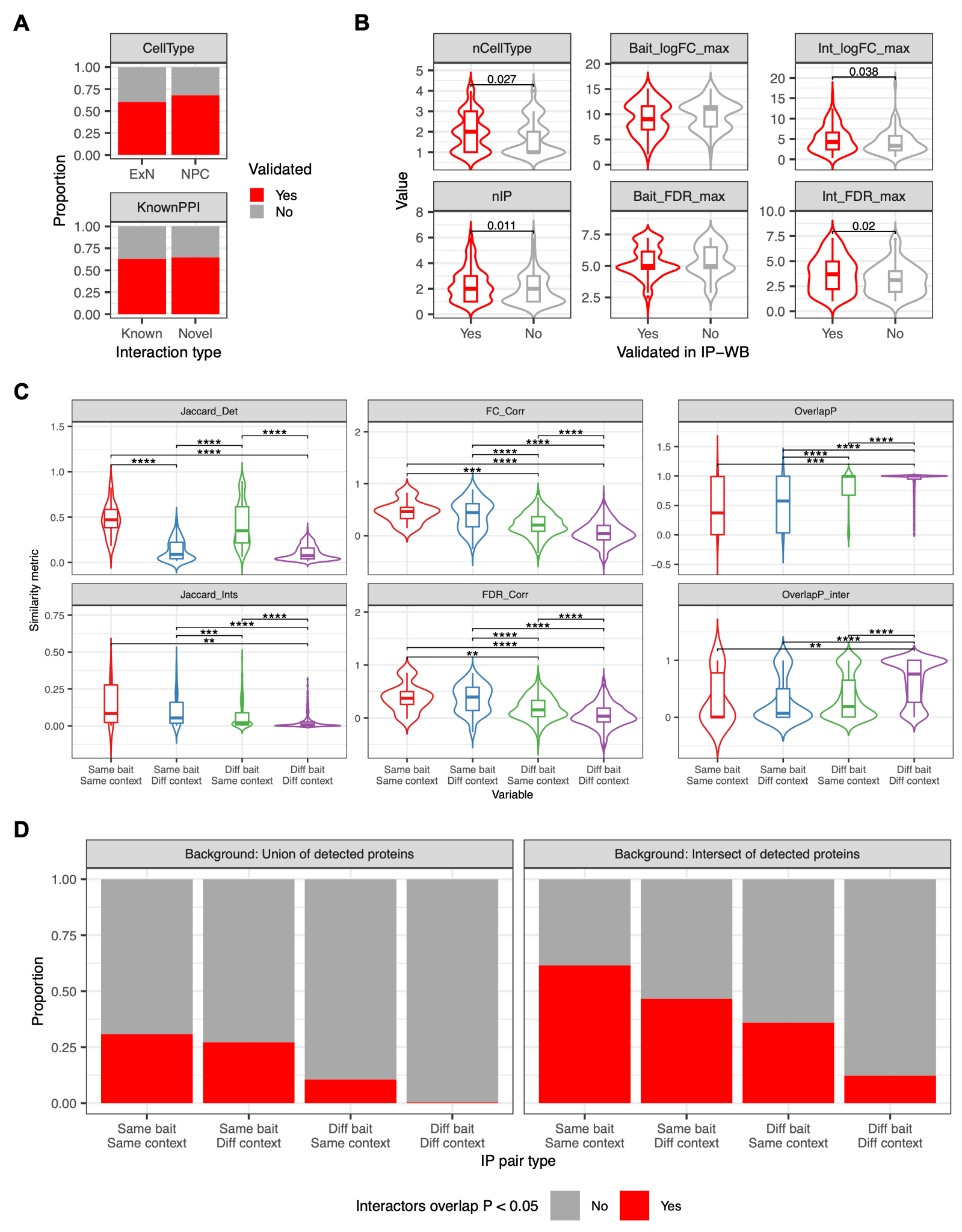

**Figure S4. Replication of identified interactions via immunoblotting and pairwise comparison of the 56 IP-MS datasets. (A)** Proportions of tested interactions validated by western blotting in independent IP experiments, stratified by cell type (top) or novelty (bottom), which was determined using PPI databases. **(B)** Characteristics of interactions validated vs. not validated by western blotting. nCellType or nIP, number of cell types or IP-MS datasets that identified the interaction; Bait_logFC_max, Bait_FDR_max, Int_logFC_max, or Int_FDR_max, maximum log_2_ FC (and corresponding -log_10_ FDR) of bait or interactor measured by IP-MS. P-values < 0.05 are shown in the plots, as calculated using two-tailed Wilcoxon rank-sum tests. **(C)** Violin and box plots showing pairwise similarity metrics for different IP-MS dataset pair types. Jaccard_Det or Jaccard_Ints, Jaccard index quantifying overlap of detected proteins or significant interactors between each pair of IPs; FC_Corr or FDR_Corr, log_2_ FC or signed (based on log_2_ FC) -log_10_ FDR correlations of proteins detected in both IPs; OverlapP or OverlapP_inter, one-tailed hypergeometric p-value indicating enrichment of interactor overlap between the IPs, using proteins detected in either IP (OverlapP) or proteins detected in both IPs (Overlap_inter) as background population. The increasing number of asterisks indicates Benjamini-Hochberg FDR < 0.05, 0.01, 0.001, or 0.0001, respectively, calculated using two-tailed Wilcoxon rank-sum tests. **(D)** Proportion of IP pairs with nominally significant interactor overlap, calculated using one-tailed hypergeometric tests and proteins detected in either IP (left) or proteins detected in both IPs (right) as background population.

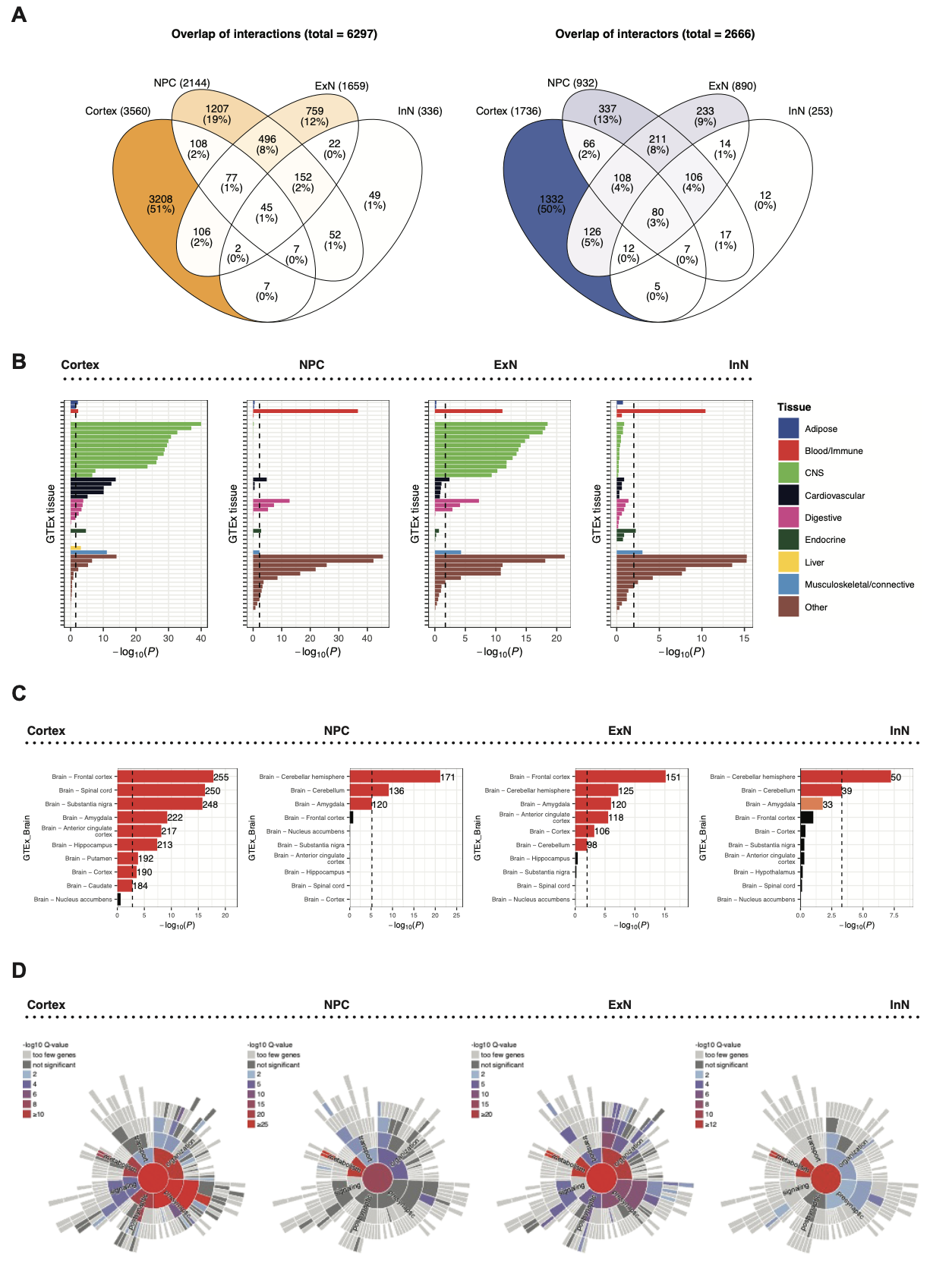

**Figure S5. Comparison of PPI networks derived from each cellular context. (A)** Overlap of PPI (i.e., bait-interactor pairs; left) or interactors (right) across the four cellular contexts. **(B)-(D)** GTEx tissue, GTEx brain region, and SynGO enrichment of the Cortex, NPC, ExN, and InN networks. Vertical dashed lines in (B) and (C) indicate FDR < 0.05. In (C), red bars indicate FDR < 0.05; orange bars indicate *P* < 0.05; number of overlapping genes (between PPI network vs. brain region-specific gene set) is shown to the right of each colored bar.

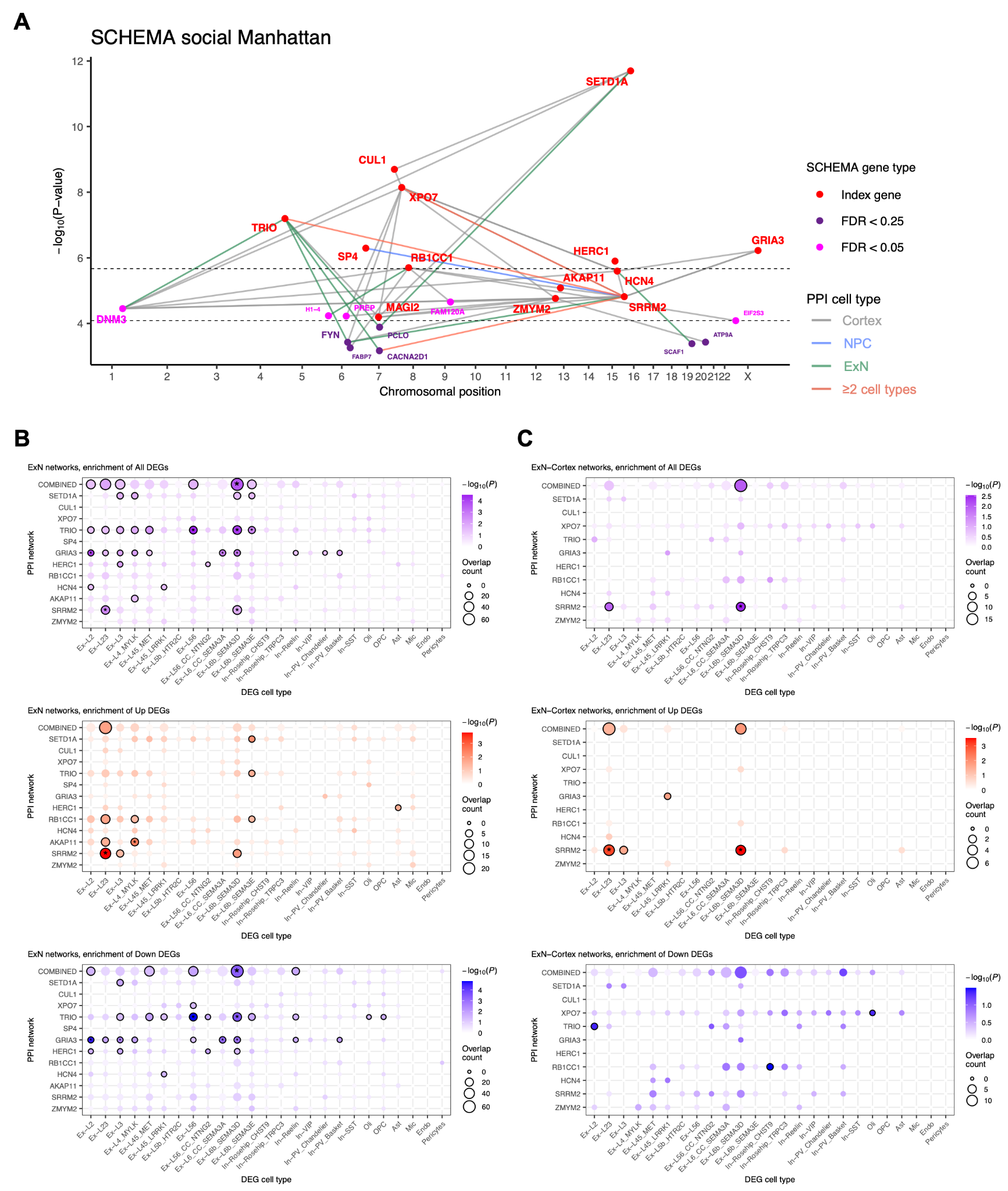

**Figure S6. Genetic and transcriptional signals in the PPI network. (A)** Social Manhattan plot showing overlap between the PPI network and genes prioritized by rare variant associations in SCHEMA. **(B)-(C)** Enrichment of cell type-specific SCZ DEGs in ExN (B) or ExN-Cortex (C) sub-networks compared against the neuronal proteome background. Black border indicates *P* < 0.05, asterisk indicates FDR < 0.05.

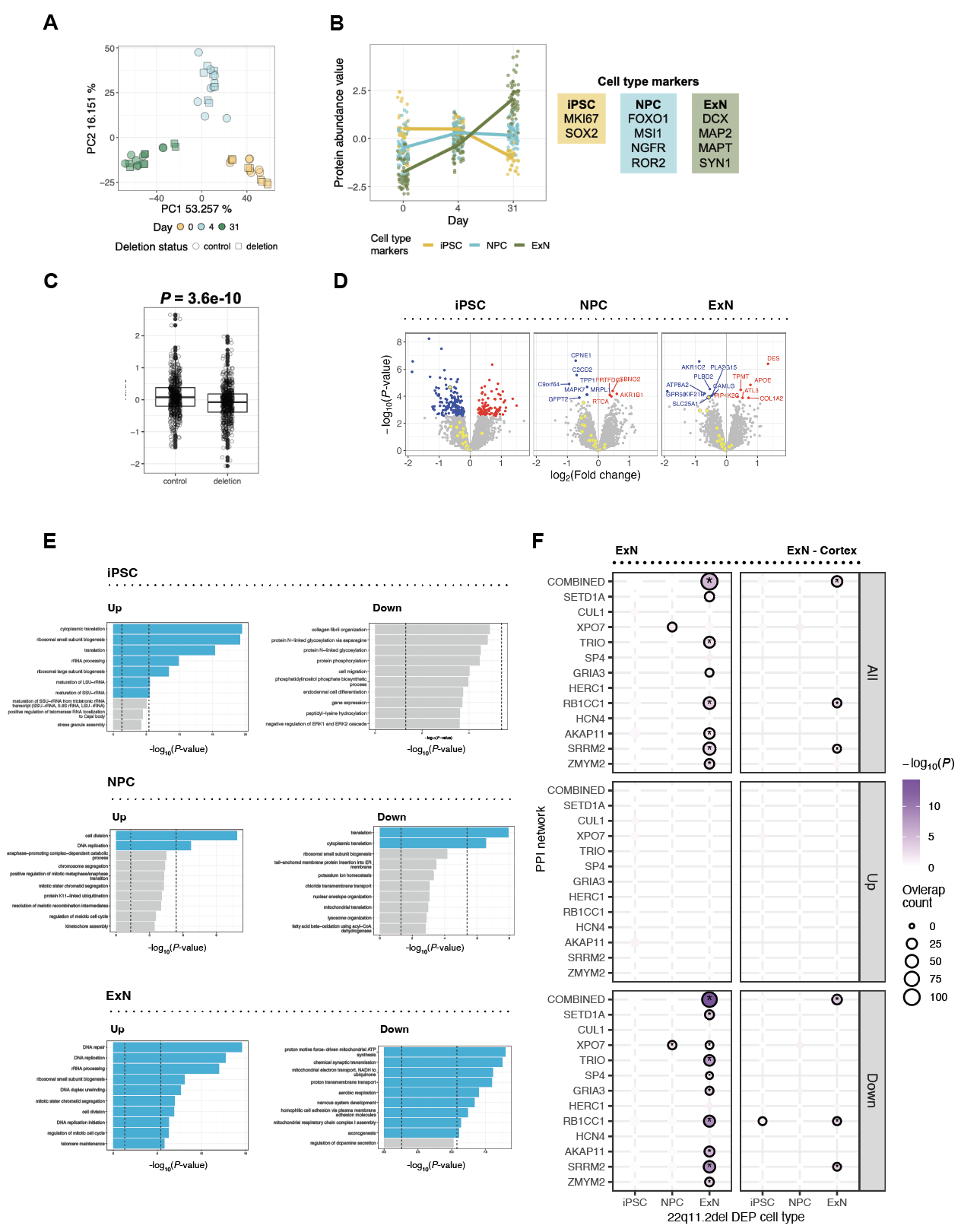

**Figure S7. Proteomic analysis of 22q11.2del vs. control cells across neuronal differentiation. (A)** Principal component analysis of proteomic profiles in 22q11.2del and control iPSC (day 0) , NPC (day 4), and ExN (day 31). **(B)** Protein expression of cell type markers in the dataset. **(C)** Expression of proteins encoded by genes in the 22q11.2del region in deletion vs. control samples; p-value was calculated using Wilcoxon rank-sum test. **(D)** Volcano plots showing differential expression of proteins in 22q11.2del vs. control iPSC, NPC, or ExN. Significant (FDR < 0.1) up or down-regulated proteins are highlighted in red or blue, respectively; 22q11.2del region proteins are highlighted in yellow. **(E)** Top enriched gene sets (GO BP terms) for DEPs in each cell state (top: iPSC, middle: NPC, bottom: ExN). Gene sets enriched in the up (left) vs. down (right) directions were determined based on the mean differential expression t-statistics of gene sets. Vertical dashed lines indicate *P* < 0.05 and *P* < 0.05/number of gene sets. **(F)** Overlap enrichment between all, up, or down-regulated DEPs (row facets) in 22q11.2 del iPSC, NPC, or ExN (x-axis) vs. the ExN or ExN-Cortex (column facets) PPI sub-networks (y-axis). Black border indicates *P* < 0.05, asterisk indicates FDR < 0.05.

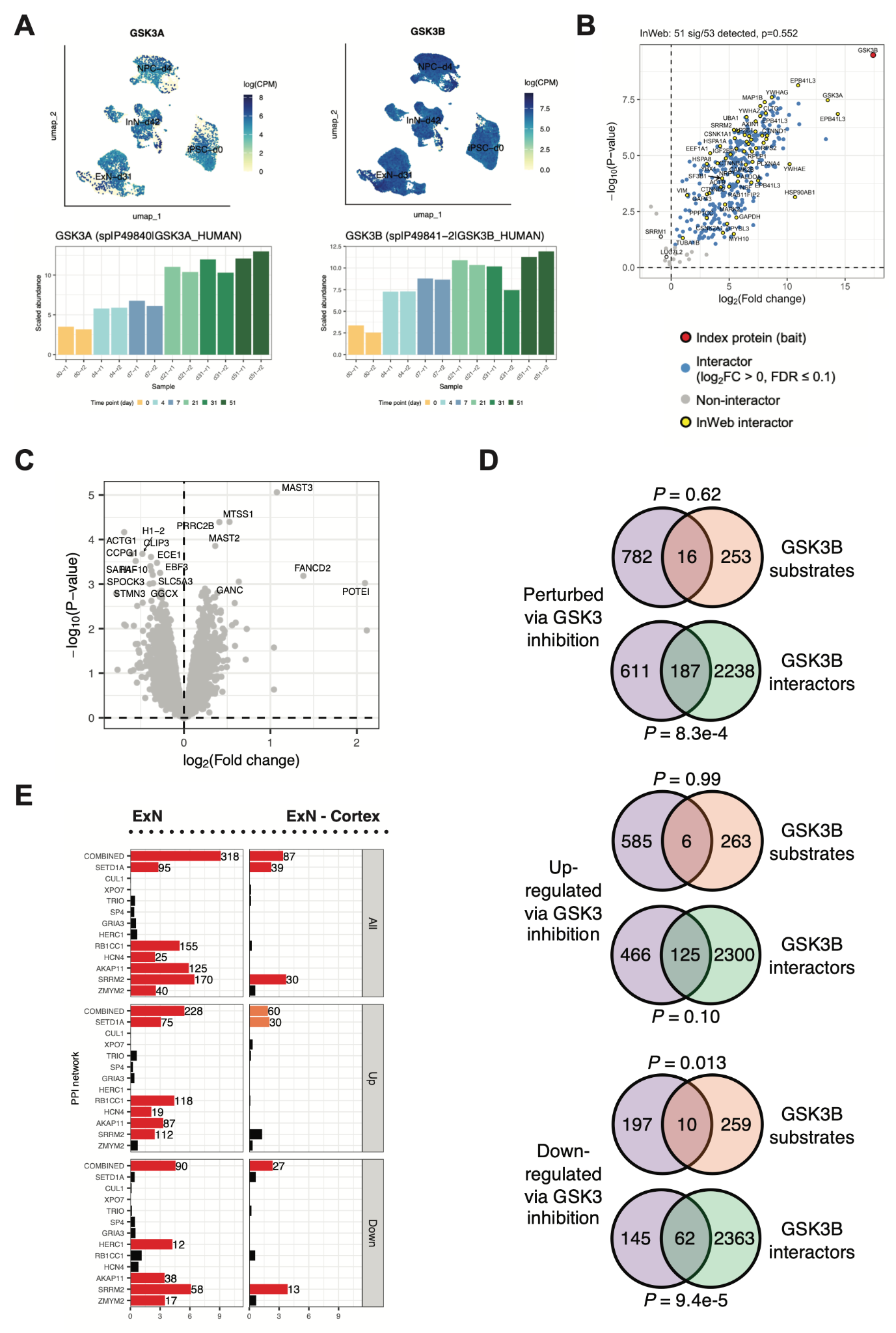

**Figure S8. Phosphoproteomic analysis of GSK3 inhibitor-treated vs. control ExN. (A)** RNA and protein expression of GSK3A and GSK3B in differentiating ExN. **(B)** Volcano plot showing IP-MS results of GSK3B in ExN. **(C)** Volcano plot showing differential expression of proteins in GSK3 inhibitor-treated vs. control ExN. No proteins were significant at FDR < 0.05. **(D)** Overlap between DE phosphopeptides vs. known GSK3B substrates or interactors. **(E)** Overlap enrichment between all, up, or down-regulated DE phosphopeptides (row facets) in inhibitor-treated ExN vs. the ExN or ExN-Cortex (column facets) PPI sub-networks. Red bars indicate FDR < 0.05; orange bars indicate *P* < 0.05; number of overlapping genes is shown to the right of each colored bar.

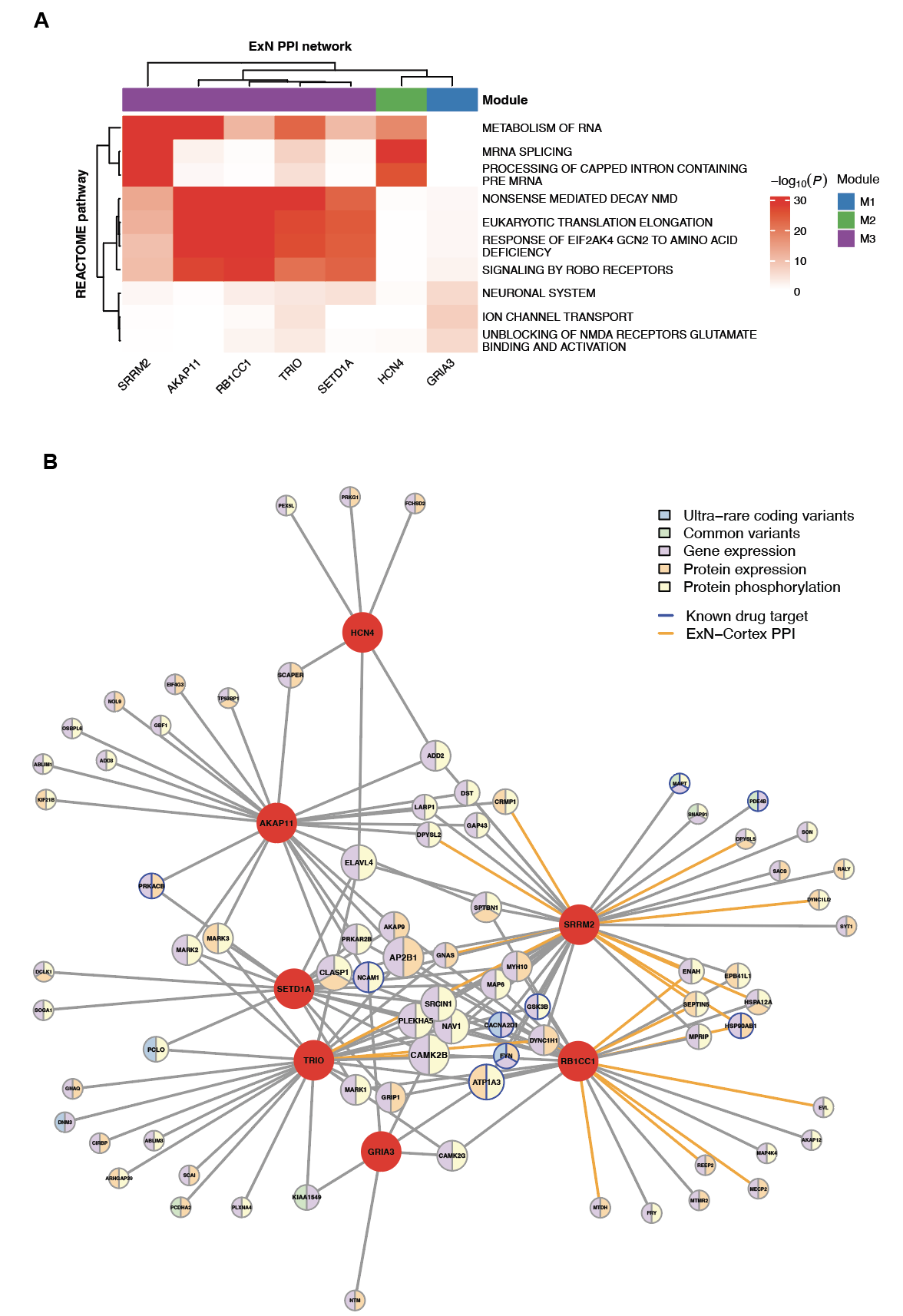
**Figure S9. Distinct SCZ modules identified through multimodal PPI network enrichment analyses**. **(A)** Clustered heat map showing REACTOME pathway enrichment patterns across the ExN sub-networks in the SCZ modules. Top three enriched pathways for each network were included. Maximum -log_10_(*P*) was capped at 30 for color visualization. **(B)** Convergent interactors perturbed in ≥2 datasets in the SCZ modules. Index proteins are shown in red; interactor proteins are color-coded based on SCZ-linked perturbation types and their sizes scale with the number of index proteins they interact with. Interactors that are known drug targets are highlighted by blue border; interactions that were identified in both ExN and cortex are highlighted in orange.
